# Long-read RNA sequencing redefines the clear cell renal cell carcinoma transcriptome and reveals novel genes and transcripts associated with disease recurrence and immune evasion

**DOI:** 10.1101/2023.09.08.23295204

**Authors:** Joshua Lee, Elizabeth A. Snell, Joanne Brown, Rosamonde E. Banks, Daniel J. Turner, Naveen S. Vasudev, Dimitris Lagos

## Abstract

**Background:** Long-read direct RNA sequencing (DRS) and PCR cDNA sequencing (PCS) of tumour samples could lead to discovery of novel transcript isoforms, novel genes, and transcriptomic co-dependencies missed by conventional short-read sequencing. However, only a handful of reports using DRS or PCS in cancer exist with no direct comparison between the two methods. Clear cell renal cell carcinoma (ccRCC) is the most common form of kidney cancer. Following primary tumour resection approximately 30% of patients experience disease recurrence. Long-read RNA sequencing has not been applied to kidney cancer.

**Methods:** 12 primary ccRCC archival tumours (discovery cohort), 6 from patients who went on to relapse, were analysed by Oxford Nanopore DRS and PCS. Results were validated in an independent cohort of 20 patients and compared to DRS analysis of RCC4 cells.

**Results:** DRS and PCS were successfully performed achieving high read length, with PCS achieving higher sequencing depth. Differentially expressed gene sets in patients who went on to relapse were determined with good overlap between DRS and PCS. Deconvolution analysis showed a loss of immune infiltrate in primary tumours of patients who relapse and revealed the CD8^+^ T cell exhaustion marker *TOX* as a novel recurrence-associated gene. Notably, novel transcript analysis revealed more than 10,000 uncharacterised candidate novel transcripts detected by both methods and in a ccRCC cell line *in vitro*. This allowed the definition of the full exonic structure of ccRCC-associated splice variants, including variants of *MVK* and *HPCAL1*. Remarkably, this also revealed a novel s*PD-L1* transcript encoding for the soluble version of the protein with a longer 3’UTR and lower stability in ccRCC cells than the annotated transcript. Levels of *sPD-L1* transcripts are unchanged in primary tumours that go on to relapse, whereas membrane *PD-L1* shows a trend towards down-regulation. Finally, both methods identified 414 novel genes, also detected in ccRCC cells *in vitro*, including a novel non-coding gene over-expressed in patients who relapse.

**Conclusions:** PCS and DRS can be used in tumour samples to uncover substantial yet unmapped features underpinning the plasticity and instability of cancer transcriptomes which are linked to disease progression and immune evasion.

## Background

Kidney cancer contributes approximately 2% of all newly diagnosed cancer cases worldwide [1]. The most common form of kidney cancer is renal cell carcinoma (RCC) and the most frequent RCC type is clear cell RCC (ccRCC, ∼75% of all RCC cases [2]). Inactivation of the *VHL* gene function is an almost universal hallmark of ccRCC. However, this is not sufficient for tumourigenesis and secondary mutations are required in hotspot genes, including *PBRM1*, *SETD2* and *BAP1*, as well as copy number changes in chromosomes 9p and 14q [3]. Of note, ccRCC tumours contain one of the highest percentages of tumour-infiltrating immune cells amongst all cancer types at approximately 30% of all cells [2, 4, 5]. Treatment of localised ccRCC typically involves removal of part or all of the kidney (radical/partial nephrectomy). Approximately one-third of patients have metastases detected at pre-operative screening and 30-50% develop metastases after the removal of the primary tumour [6]. Several approaches have been proposed for assessing the risk of disease recurrence following surgery [7]. These are often based on pathological and clinical features, such as the Tumour-Node-Metastasis (TNM) staging system [8], the Leibovich score [9], and other similar scores [10, 11]. Scores based on gene expression signatures have also been proposed to refine risk prediction [12–14]. However, despite a recognised need [15, 16], so far, no set of biomarkers has reached routine clinical practice.

Aberrant co– and post-transcriptional events (e.g. alternative splicing/polyadenylation, post-transcriptional modifications, etc), drive oncogenesis but also tumour immunogenicity [17, 18]. Our understanding of cancer transcriptomes is nearly exclusively based on short read sequencing platforms. Given that the average length of a mRNA is 1.5-2kb in mammals this approach requires high depth of sequencing to confidently call transcript variants and is limited with regards to reconstruction of full-length novel transcripts. Often the reliance on reference genomes/transcriptomes means that this approach misses or discards novel transcripts. Furthermore, it is extremely difficult, if not impossible, to confidently establish transcriptional co-dependencies, i.e. co-existence of distinct features (e.g. specific splice junctions and untranslated regions – UTRs) on the same transcript. Long-read direct RNA sequencing (DRS) and PCR cDNA sequencing (PCS) have emerged as transformative methodological alternatives to overcome these limitations [19]. Yet, in cancer, there are only a handful of reports using long read sequencing in tumour samples from patients with solid [20–22] or blood cancers [23–25], with only one example of using DRS (in 3 myeloma patient samples [24]). Currently, there are no reports directly comparing PCS to DRS in clinical samples and long-read sequencing has not been applied to kidney cancer.

Here, we successfully employed Nanopore DRS and PCS [26] to explore ccRCC transcriptomes using 2 μg and 200 ng total RNA, respectively, from archival surgical fresh frozen specimens. Comparing patients who went on to experience recurrence against controls generated a list of differentially expressed genes (DEGs) with good overlap between the two methods. Deconvolution analysis revealed a loss of immune infiltrate and specifically CD8^+^ T cells in primary tumours of patients who relapse. Both methods identified thousands of novel isoforms of known genes and 414 novel genes, all of which were also detected by DRS in the ccRCC cell line RCC4, indicating cancer cell-intrinsic expression. Novel transcript analysis allowed us to determine the full exonic structure of ccRCC-associated splice variants (SVs). Targeted analysis of immune checkpoint transcripts revealed a novel *PD-L1* transcript encoding for the soluble version of the protein but having a longer 3’UTR than the currently annotated transcript. The *sPD-L1* and *mPD-L1* transcripts show differential stability and responsiveness to inflammatory cytokines *in vitro*. Finally, we report the discovery of a novel non-coding gene, over-expressed within primary tumours of patients who go on to experience recurrence. Overall, our study uncovers a substantial new layer of in ccRCC and specific features associated with disease recurrence and immune evasion.

## Methods

### Study participants and ethics

In this observational study, we used 32 ccRCC tumour nephrectomy samples (16 non-recurrent and 16 recurrent cases) collected between 2000 and 2012 and stored in the Leeds multidisciplinary research tissue bank (regional ethics committee approval: Yorkshire & The Humber – Leeds East Research Ethics Committee, reference 15/YH/0080). 12 samples were used as a discovery cohort for DRS and PCS sequencing (6 non-recurrent and 6 recurrent) and 20 (10 in each group) were used as an independent validation cohort. For the recurrence group, median time to relapse was 23 months (5 – 176). For the control group median follow-up without relapse was 11 years (7 – 18). Groups were matched for demographic, pathological, and clinical characteristics including TNM and Leibovich score (**Additional file 1: Table S1**, age is shown in 5-year intervals). The sample/kidney IDs were not known to anyone outside the research group.

### Tissue sample preparation

Following surgical removal, tissue samples were washed in phosphate-buffered saline (PBS), blotted on a tissue before being enveloped in aluminium foil and snap frozen in liquid nitrogen. Once thawed, samples were immediately used for RNA extraction without further freeze-thawed cycles. All cases underwent pathology review of a parallel formalin fixed paraffin embedded (FFPE) block to confirm ccRCC histology and tumour cell viability, as part of a separate study [27].

### Cell culture and cytokine treatment

RCC4 cells were maintained at 37°C in a humidified atmosphere of 5% CO_2_ and grown in complete Dulbecco’s Modified Eagle’s Medium (DMEM, Gibco 21969-05), supplemented with 10% foetal bovine serum (FCS) (Gibco A5256701), 1% 200 mM L-Glutamine (Gibco 25030) and 1% penicillin/streptomycin (Gibco 15140). For RCC4 Direct RNA sequencing experiment, 1 x 10^6^ RCC4 cells were seeded in 15 mL of complete DMEM in T75 flasks. 24 hours after seeding, media were changed into complete DMEM, with or without the addition of IFN-γ (1000U/mL, Peprotech 300-02) and TNF (25 ng/mL, Peprotech 300-01). Cells were harvested 24 hours later for RNA extraction. Three flasks of T75s were used for each replicate for the sequencing experiment.

### RNA extraction

Total RNA was extracted from nephrectomy specimens or cultured cells using QIAzol (Qiagen 79306) and RNeasy kits (Qiagen 74004) with on-column DNase I digestion step, according to manufacturer’s instruction. Nephrectomy specimens were homogenized in QIAzol using a TissueLyser LT (Qiagen 85600) with stainless steel beads (Qiagen 69997). RNA integrity number (RIN) was determined using the 2100 Bioanalyzer with RNA Nano kit (Agilent 5067) and quantified using Qubit RNA HS assay kit (Invitrogen, Q32852). Total RNA from RCC4 for Direct RNA sequencing was enriched for Poly(A)^+^ RNA molecules using the Dynabeads Oligo(dT)_25_ mRNA isolation kit (Invitrogen 61002).

### Library preparation and RNA sequencing

Sequencing libraries used for PCR-cDNAseq and Direct RNAseq were generated using the SQK-PCS111 and SQK-RNA002 kit (Oxford Nanopore Technologies, ONT), respectively. For the nephrectomy specimens, 200 ng and 2 μg of extract total RNA were used as input for each sequencing library for PCR-cDNAseq and Direct RNAseq, respectively. 500 ng of poly(A)^+^ RNA was used for each sequencing library for Direct RNAseq of RCC4 cells. For PCR-cDNAseq, cDNA libraries were prepared with the SQK-PCS111 kit according to manufacturer’s instruction with 14 cycles of PCR cycles. For Direct RNAseq, libraries were prepared with the SQK-RNA002 kit according to manufacturer’s instruction including the optional reverse transcriptase step. Sequencing libraries for each experiment were prepared together to mitigate batch effects. All sequencing libraries were sequenced on ONT PromethION sequencer with R9.4.1 PromethION flow cells (ONT) for 72 hours. Basecalling and FASTQ files generation were performed with Guppy (version 5.1.12, ONT).

### Quality control and reads alignment

Sequencing reads generated from Direct RNAseq, and PCR-cDNAseq with a minimum read quality score (Q score) of 7 were used for mapping and downstream analysis. FASTQ files generated from sequencing runs were concatenated using catfishq (version 1.4.0, https://github.com/philres/catfishq). PCR-cDNAseq reads were orientated by pychopper (version 2.5.0, https://github.com/nanoporetech/pychopper), filtered for the presence of 5’ and 3’ sequencing adaptors and trimmed by cutadapt (version 4.1) [28]. Direct RNAseq and processed PCR-cDNAseq reads were aligned to either human genome, transcriptome (GRCh38, Ensembl release 105) or Stringtie assembly using minimap2 (version 2.24), with recommended parameters (Genome alignment: –ax splice –uf –k14; Transcriptome alignment:-ax map-ont –p 0 –N 10) [29]. Aligned reads were sorted, merged and indexed to BAM files with samtools (version 1.13) [30]. The workflows for reads alignment are available at https://github.com/joshuacylee/DRS and https://github.com/joshuacylee/PCR-cDNAseq. Mapping data quality and statistics of sequencing data were analysed by Nanoplot and bamslam [31], https://github.com/josiegleeson/BamSlam).

### Differential gene expression

Gene-level expression quantification was performed using featureCounts with long-read counting mode (-L) (subread version 2.0.0) [32]. Transcript isoform quantification was performed using NanoCount (version [33]. Normalisation and identification of differentially expressed genes (p_adj_ ≤ 0.1 and log_2_FoldChange ≥ 2) were performed using the R package DESeq2 (version 1.40.2) [34]. PCA plots were generated by DESeq2 and volcano plots were generated with the R package EnhancedVolcano (version 1.18.0). Workflow for differential gene expression identification is available at (https://github.com/joshuacylee/DESeq2).

### Gene set enrichment analysis and tumour-infiltrating immune cell analysis

Gene set enrichment analysis was performed using clusterProfiler (v4.4.4) [35]. Gene ontology biological process and molecular function databases were used for functional enrichment analysis. Parameters used for gene ontology enrichment were as follows: Permutations (nPerm):10000, minimum gene set size (minGSSize): 5, Maximum gene set size (maxGSSize): 500, Minimum p-value (pvalueCutoff) = 0.05, Organism (Orgdb) = org.Hs.eg.db, pAdjustMethod = Benjamini-Hochberg (BH). Tumour purity and tumour-infiltrating immune cell population abundance was estimated using two gene signature-based algorithms: ESTIMATE(v1.0.13) and xCell (v1.1.0) [4, 36]. Tumour-infiltrating immune cell type deconvolution was performed using CIBERSORTx and EPIC [37, 38].

### Transcriptome assembly and novel gene/isoform discovery

Using reference genome aligned PCR-cDNAseq bam files, transcript assembly was performed with StringTie2 and FLAIR [39]. StringTie2 assembly was performed with long-reads processing mode (-L), guided by reference gene annotation (Ensembl release 105).StringTie2 transcriptome assemblies from all sequenced nephrectomy specimens were then merged using the –merge option to generate transcript annotation file (gtf file). StringTie2 annotation used for novel gene mapping was performed by merging all nephrectomy assemblies with reference gene annotation (Ensembl release 105). FLAIR assembly was generated using the ‘flair correct’ and ‘flair collapse’ commands, with the long-read optimised option selected (--trust_ends). Generated transcript annotation files from StringTie2 and FLAIR were compared to Ensembl reference gene annotation (with –r option) where each assembled transcript was classified with a classcode using GffCompare (v0.12.6) [40]. In accordance with [33], transcripts were categorised into 3 main categories: ‘Known’ (‘=’: Complete intron chain match, ‘c’: Partial intron chain match), ‘Novel’ (‘j’: Multi-exon with at least 1 matched junction’, ‘k’: Containing reference, ‘m’: Retained intron(s) – all covered, ‘n’: Retained intron(s) – not all covered, ‘i’: Contained within intron, ‘o’: Overlapped exon, ‘x’: Overlapped antisense, ‘y’: Containing reference within intron, ‘u’: None of above/Unknown), and ‘Potential Artefacts’ (‘p’: No over-lap, ‘e’: Single exon partially covering an intron, ‘s’: Intron matched on opposite strand, ‘r’: Repeat).

StringTie2 assembled transcripts were also characterised by SQANTI3 (v5.1.2), which classifies genes as ‘annotated’ or ‘novel’, and isoforms as full splice match (FSM), incomplete splice match (ISM), novel in catalog (NIC), novel not in catalog (NNC), antisense, genic intron, genic genomic and intergenic [41]. FSM represent isoforms with the exact same splice junctions and number of exons with the reference annotation. ISM represent isoforms with fewer exons from the 5’ end but with the remaining internal splice junction sites matching with the reference annotation. NIC isoforms contain novel combinations of known splice junctions/exons compared with the reference annotation. NNC represents isoforms with at least one novel, unannotated splice site. In the SQANTI3 model, FSM and ISM represent the ‘Known’ transcripts, whereas NIC, NNC, antisense, genic intron, genic genomic and intergenic isoforms represent the ‘Novel’ transcripts. SQANTI3 also predicts coding potential of transcripts using the GeneMarkS-T model [42]. Integrative genomics viewer (IGV) tracks and reference genome mapped reads aligned to the region of novel genes and isoforms were visualized using IGV viewer [43].

### cDNA synthesis and qPCR

RNA molecules were reverse transcribed to cDNA molecules using oligo(dT) primer (Novagen 69896) and SuperScriptII reverse transcriptase (Invitrogen 18064022). qPCR assays were performed using Fast SYBR Green master mix (Applied Biosystem 4385612) and pre-validated primers (Eurofins) on a StepOnePlus real-Time PCR system (Applied Biosystem) for 40 amplification cycles. Relative transcript levels were determined using the ΔΔCt (cycle threshold) method with *GAPDH* and *ACTB* used as loading controls. Details on the primers used can be found in **Additional file 1: Table S2**.

### RNA stability assay

4 x 10^4^ RCC4 cells were seeded in 12 well plates. 24 hours after seeding, media were changed into complete DMEM, with or without the addition of IFN-γ (1000U/mL) and TNF (25 ng/mL). 24 hours later, Actinomycin D (2 μg/mL, Generon) was added and cells were harvested after 0, 2, 4, and 8 hours of incubation for RNA extraction and qPCR. 3 wells were used as technical replicates for each biological replicate (n = 3) for each time point.

### Statistical analysis

Statistical analysis was performed using GraphPad Prism 9. two-tailed Mann-Whitney U tests were used to compare non-parametric analysis of gene or transcript isoform expression levels, tumour purity estimations, immune scores and relative immune cell populations between experimental groups, with p ≤ 0.05 considered statistically significant. For comparison of more than two groups, Kruskal-Wallis test was used with p ≤ 0.05 considered significant. For correlative analysis, R^2^ (coefficient of determination) was used to calculate the goodness of fit between datasets, and P values were generated from F-test, with p ≤ 0.05 considered statistically significant. Differential gene expression analysis by DESeq2 implements the Wald test, followed by false discovery rate correction by the Benjamini-Hochberg Method. Genes with p_adj_ < 0.05 and |log_2_FoldChange| ≥ 2 are considered to be significantly differentially expressed. All p values of non-significant results are indicated in graphs.

## Results

To demonstrate the feasibility of utilising long-read RNA sequencing technologies for characterising ccRCC transcriptomes in archival surgical fresh frozen specimens, we sequenced twelve snap-frozen nephrectomy samples using Nanopore PCS and DRS on ONT PromethION flow cells. These samples consisted of six specimens from patients who later developed ccRCC recurrence and six non-recurrent controls (see methods and **Additional file 1: Table S1**). An overview of study design and data analysis pipeline is shown in **Fig. 1A**.

**Figure 1:**
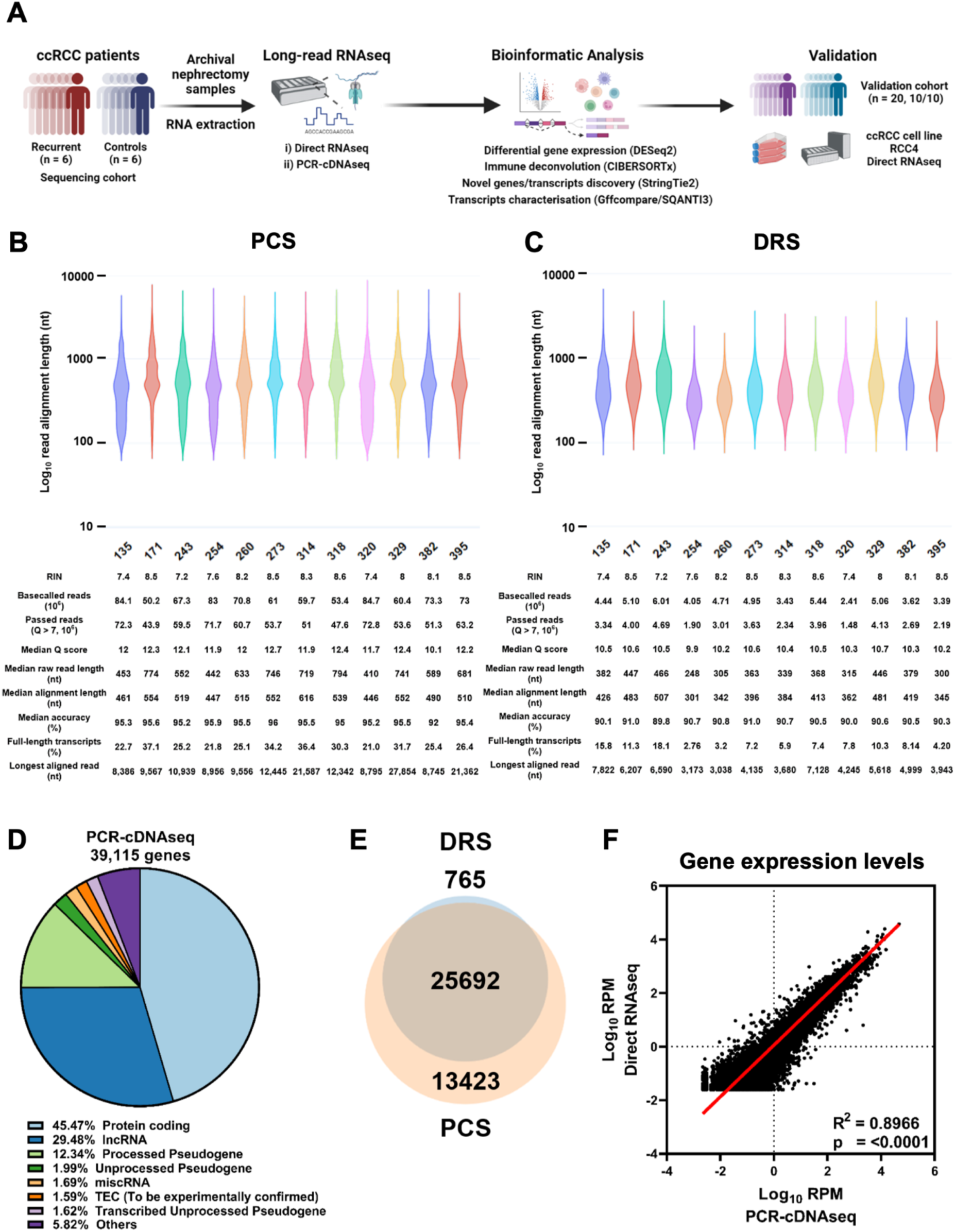
DRS and PCS of ccRCC nephrectomy samples. **A.** Summary of study design and data analysis workflow – figure made with Biorender. **B.** Violin plot showing Log_10_ transformed raw read lengths of passed reads generated by PCS. RIN score, number of basecalled reads, number of passed reads (Q >7), median read Q score, median raw read length (nt), median reference genome (Ensembl release 105, GRCh38) alignment length (nt), median read accuracy, percentage of reads representing full-length transcripts (95%+ coverage of reference transcript isoform) and the length of the longest reference genome aligned read for each sequencing dataset are listed in the table below violin graph. **C.** as in **B**, but for DRS. **D.** Pie chart depicting the proportions of gene biotypes of all mapped genes from reference genome (Ensembl release 105, GRCh38) mapped PCS reads of sequenced tumour samples. **E.** Venn diagram showing the overlap between PCS and DRS mapped genes. **F.** Correlation between gene expression levels (Log_10_ Reads per million (RPM)) of all genes mapped by both PCS and DRS (n = 25,692). Diagonal line represents the line of best fit. R^2^ value was computed to measure goodness-of-fit and p value was generated from F-test, with p<0.05 considered statistically significant.

### DRS and PCS of ccRCC nephrectomy samples

All nephrectomy specimens were successfully sequenced using both PCS and DRS. After 72 hours of sequencing, PCS generated reads ranging from 50 million to 85 million (median = 56.6 million, total = 701 million), with approximately 80% qualified as pass reads (median = 45.8 million, total = 561 million) having a minimum read Q score of 7. DRS generated between 2.4 million to 5.5 million reads (median = 4.6 million, total = 52.6 million), with approximately 70% qualified as pass reads (median = 3.2 million, total = 37.4 million). Summary sequencing output statistics can be found in **Additional File 1: Table S3**.

Both PCS and DRS reads were next mapped to the human reference genome and transcriptome. The median alignment length for PCS and DRS reads were 517 and 405 nucleotides respectively. A range of 21 – 37.1% (median = 25.95) of PCS– and 3.2 – 18.1% (median = 7.6%) of DRS-aligned reads represent full-length transcripts (coverage of at least 95% of the mapped reference annotation) (**Fig. 1B, C**). Overall, PCS and DRS reads achieved median accuracies of 95.5% and 90.5%. The longest aligned reads for PCS and DRS were 27,854 and 7,822 nucleotides, respectively. These results demonstrated the capability of Nanopore long-read RNA sequencing to produce high-depth, long-read sequencing datasets from flash-frozen, archival clinical specimens.

To evaluate the ability of long-read sequencing to capture the diversity of the transcriptome, we examined the RNA biotypes of genes identified by PCS and DRS. PCS identified 39,115 genes across the 12 samples, with a median of 26,203 mapped genes per specimen (**Fig. S1A**). Among all PCS-mapped genes, 45.47% were classified as protein-coding genes, 29.48% as long non-coding RNAs (lncRNA) and 12.34% as processed pseudogenes (**Fig. 1D**). In comparison, DRS identified 26,457 genes across the specimens (median = 18,057 per sample), with 77.69% classified as protein-coding genes, 15.22% as lncRNAs and 3.07% as processed pseudogenes (**Fig. S1B**). 25,692 genes were mapped by both methods (**Fig. 1E**). 13,423 genes were exclusively mapped by PCS, likely due to higher sequencing depth compared to DRS. Notably, 765 genes were exclusively mapped by DRS.

We observed that the majority of expressed genes for both PCS and DRS were protein-coding (89.4% and 91.7%, respectively), followed by mitochondrial rRNAs (mt-rRNAs) (5.35% and 4.66%), processed pseudogenes (2.57% and 1.74%) and lncRNA (1.71% and 1.10%) (**Fig. S1C**). Interestingly, despite using total RNA as input for PCS and DRS library preparation without applying poly(A)^+^ enrichment or ribo-depletion methods, highly abundant rRNAs were sequenced at negligible levels. Distribution of gene expression levels for each biotype by PCS and DRS are illustrated by violin plots at **Fig. S1D**. Furthermore, amongst genes mapped by both PCS and DRS (n = 25,692), we found significant correlation in their gene expression levels (**Fig. 1F** and **S2**).

Overall, whilst PCS provided greater sequencing depth, our data demonstrated that both methods can capture a diverse range of transcripts from archival clinical samples, yielding highly concordant gene expression profiles.

### Differential gene expression analysis reveals that ccRCC recurrence is associated with suppressed tumour immune infiltration

We then tested whether DRS and PCS can identify features associated with ccRCC recurrence. When including the whole transcriptome, PCA did not result in visually distinct clusters correlating with ccRCC recurrence status for either PCS or DRS (**Fig. S3A**). No sample separation was observed based on RNA integrity (**Fig. S3B**). However, differential gene expression analysis identified 136 and 34 genes with significantly differential expression (|log_2_FoldChange| ≥ 2, p_adj_ ≤ 0.1) between recurrent and non-recurrent tumours by PCS and DRS, respectively, with substantial overlap (**Fig. 2A, B, Additional File 1: Table S4 and S5**). The directionality of gene expression amongst these differentially expressed genes (DEGs) showed strong correlation (**Fig. 2C**).

**Figure 2:**
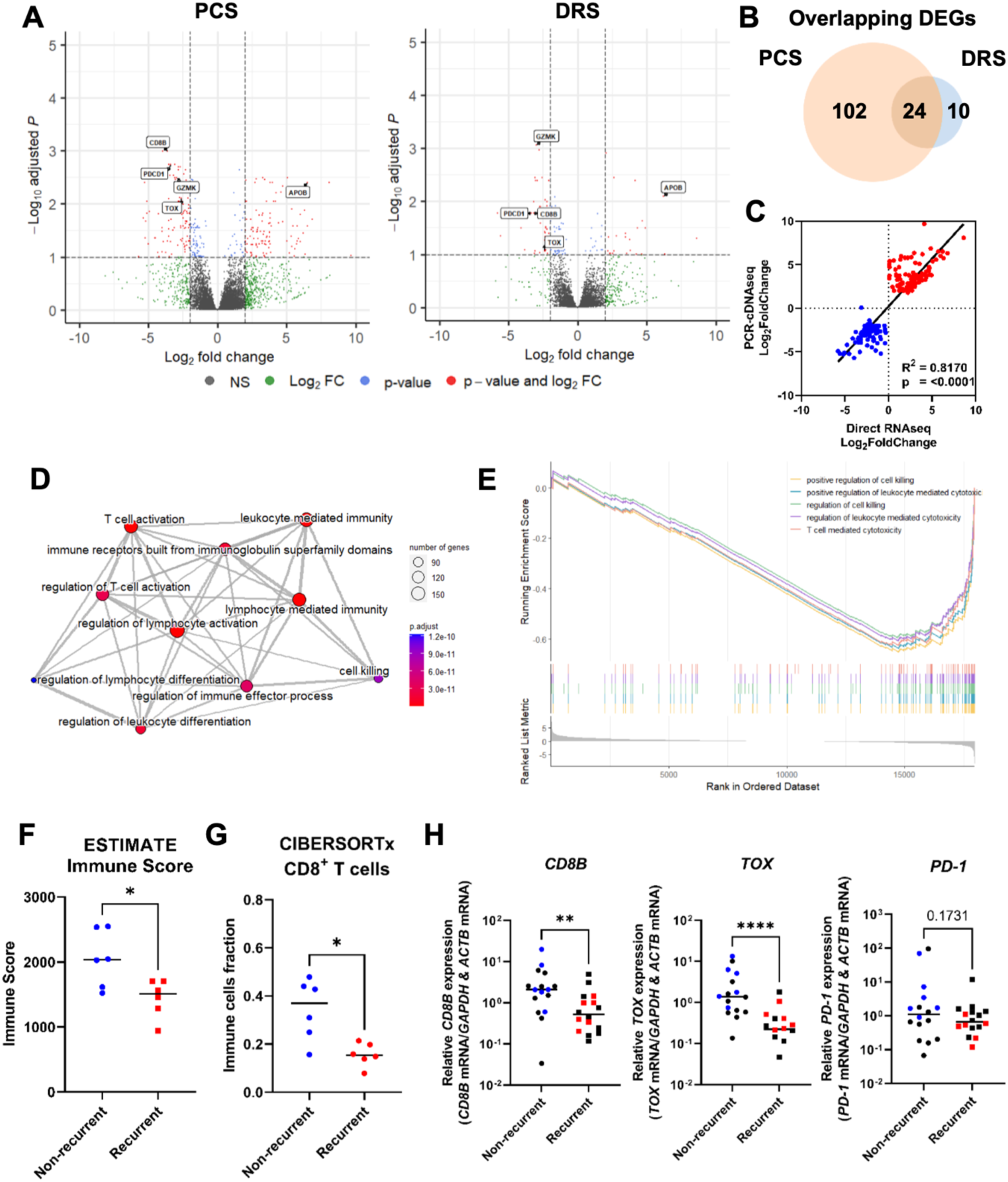
ccRCC recurrence is associated with suppressed tumour immune infiltration. **A**. Volcano plots showing differentially expressed genes (red) between recurrent and non-recurrent ccRCC tumours from PCS and DRS data using Ensembl gene reference (Ensembl release 105). Dotted lines indicate significance threshold (|log_2_FoldChange| ≥ 2, p_adj_ ≤ 0.1). Names of genes that were validated by qPCR with validation cohort are shown. **B.** Venn diagram showing overlaps of DEGs identified by both PCS and DRS. **C.** Correlation between log_2_FoldChange of DEGs identified by either or both PCS and DRS (recurrent vs non-recurrent ccRCC tumours). Blue and red dots represent significantly down– and up-regulated genes by either or both PCS and DRS. Diagonal line represent the line of best fit. R^2^ value was computed to measure goodness-of-fit and p value was generated from F-test, with p ≤ 0.05 considered statistically significant. **D.** Gene Ontology Biological Process GSEA map showing top 10 enriched terms associated with ccRCC recurrence-associated differential gene expression by PCS. **E.** GSEA enrichment plot for the top 5 enriched Gene Ontology Biological Process terms associated with ccRCC recurrence differential gene expression from DRS. The x-axis shows genes represented in each pathway and the y-axis shows enrichment scores. **F.** Grouped dot plot showing estimated immune score of non-recurrent (blue) and recurrent (red) ccRCC tumour by the ESTIMATE algorithm, using PCS gene expression data. **G.** Grouped dot plot showing relative population of CD8^+^ T cells within immune infiltrates of non-recurrent (blue) and recurrent (red) ccRCC tumours estimated by CIBERSORTx using PCS gene expression data. **H.** *CD8B*, *TOX* and *PD-1* mRNA levels measured by qRT-PCR in recurrent and non-recurrent tumours from sequenced cohort (blue and red, n = 12) and validation cohort (black, n = 20), relative to average mRNA levels in non-recurrent tumours. mRNA levels were normalised to *GAPDH* and *ACTB*. For **F – H**, two-tailed Mann-Whitney U tests were used with p ≤ 0.05 considered significant. * = p < 0.05, ** = p < 0.01, **** = p < 0.0001. p value of non-significant results is indicated in graph. Centre line represents median for each group.

Within the overlapping DEGs between PCS and DRS, several key adaptive immune genes, including *CD8B*, *PDCD1*, *GZMK* and *TOX*, were significantly downregulated in recurrent samples (**Fig. 2A, B**). To evaluate variations in biological processes and pathways between recurrent and non-recurrent ccRCC tumours, we performed gene set enrichment analysis (GSEA). Using the gene ontology (GO) biological processes (BP) database, we found that the top 10 most significantly enriched (by p_adj_) GO BP terms from PCS data were all associated with adaptive immunity (**Fig. 2D**). This pattern was also found using DRS data, where enrichment plot for the top 5 enriched GO BP terms (by p_adj_) by DRS showed pronounced suppression of adaptive immune response related pathways (i.e. positive regulation of cell killing, T cell mediated cytotoxicity) in ccRCC recurrent samples (**Fig. 2E**).

To further explore the relationship between ccRCC recurrence and immune infiltrate populations, we used the ESTIMATE algorithm, which uses gene expression signatures to infer tumour purity and immune cell abundance. Using PCS data, we found that recurrent ccRCC exhibited significantly lower immune scores and higher levels of tumour purity compared with non-recurrent controls (**Fig. 2F, S4A**). DRS data displayed a borderline non-significant trend towards decrease in immune scores in recurrent ccRCC tumours (p = 0.0881) and a high degree of concordance was found between DRS and PCS ESTIMATE immune scores (R^2^ = 0.87, p < 0.0001) (**Fig. S4B,C**). Both PCS and DRS datasets also had significantly lower immune scores for recurrent samples according to xCell analysis (**Fig. S4D**).

Next, to further evaluate our findings, the immune infiltrate profiles were further analysed using another cell-type deconvolution algorithm, CIBERSORTx [37]. Both PCS and DRS exhibited a significant reduction in the fraction of CD8^+^ T cell within recurrent ccRCC tumours when compared to non-recurrent controls (**Fig. 2G**, **S4E, F**). Similarly, EPIC, another immune cell type deconvolution method, also indicated suppression of CD8^+^ T cell within the immune infiltrates amongst the recurrent ccRCC tumours (**Fig. S4G**).

These findings agreed with a previously reported, qRT-PCR based, ccRCC recurrence prediction assay, which also linked lower expression levels of immune response genes with an increased likelihood of disease recurrence [14]. Amongst the 11 recurrence-related gene makers examined in that study, our PCS and DRS analyses also identified that levels of *NOS3* and *CCL5* were significantly decreased in the recurrent tumours (**Fig. S4H**).

Critically, we sought to validate our long-read sequencing results by an independent method (qRT-PCR) and using samples from both the sequenced cohort but also an independent validation cohort (n = 20, 10 from recurrent ccRCC patients and 10 non-recurrent controls). This analysis confirmed significant downregulation of the key CD8^+^ T cell marker *CD8B* and the T cell exhaustion marker *TOX* in the recurrent tumours (**Fig. 2H, S5A, B**). Interestingly, the decreased expression of *GZMK* and *PD-1* in the recurrent tumours observed in the sequenced cohort were also validated using qRT-PCR, but the suppression was not replicated in the independent validation cohort (**Fig. 2H, S5C, D**).

Collectively, these findings demonstrated that both PCS and DRS can identify robust differential expression signatures between different classes of clinical samples. In the context of ccRCC, PCS and DRS suggested a significant suppression of immune infiltrates, particularly CD8^+^ T cells, in the tumour microenvironment of ccRCC patients who later develop disease recurrence, and identified the exhaustion marker *TOX* as a novel recurrence-associated gene.

### Long-read RNA sequencing enables the discovery of novel full-length transcripts expressed in ccRCC cells

A unique strength of long-read sequencing is the potential to discover novel transcript isoforms and genes, not currently included in the reference transcriptome. To identify novel transcript isoforms that are present in the ccRCC nephrectomy specimens, we applied StringTie2 to perform transcriptome assembly using PCS reads aligned to the reference genome. StringTie2 assembled isoforms were subsequently compared to the reference annotation (Ensembl release 105) with both SQANTI3 and gffcompare (see Methods). SQANTI3 classifies each assembled isoform into known or novel based on their splice junction matches. Known transcripts comprise FSM and ISM, whereas NIC, NNC, antisense, fusion, genic, genic intron and intergenic isoforms are classified as novel transcripts (**Fig. 3A, Additional file 1: Table S6**). Similarly, gffcompare assigns each StringTie2 assembled isoform with a transcript class code which corresponds to ‘Known’ and ‘Novel’ transcripts (**Fig. S6, Additional file 1: Table S6**).

**Figure 3:**
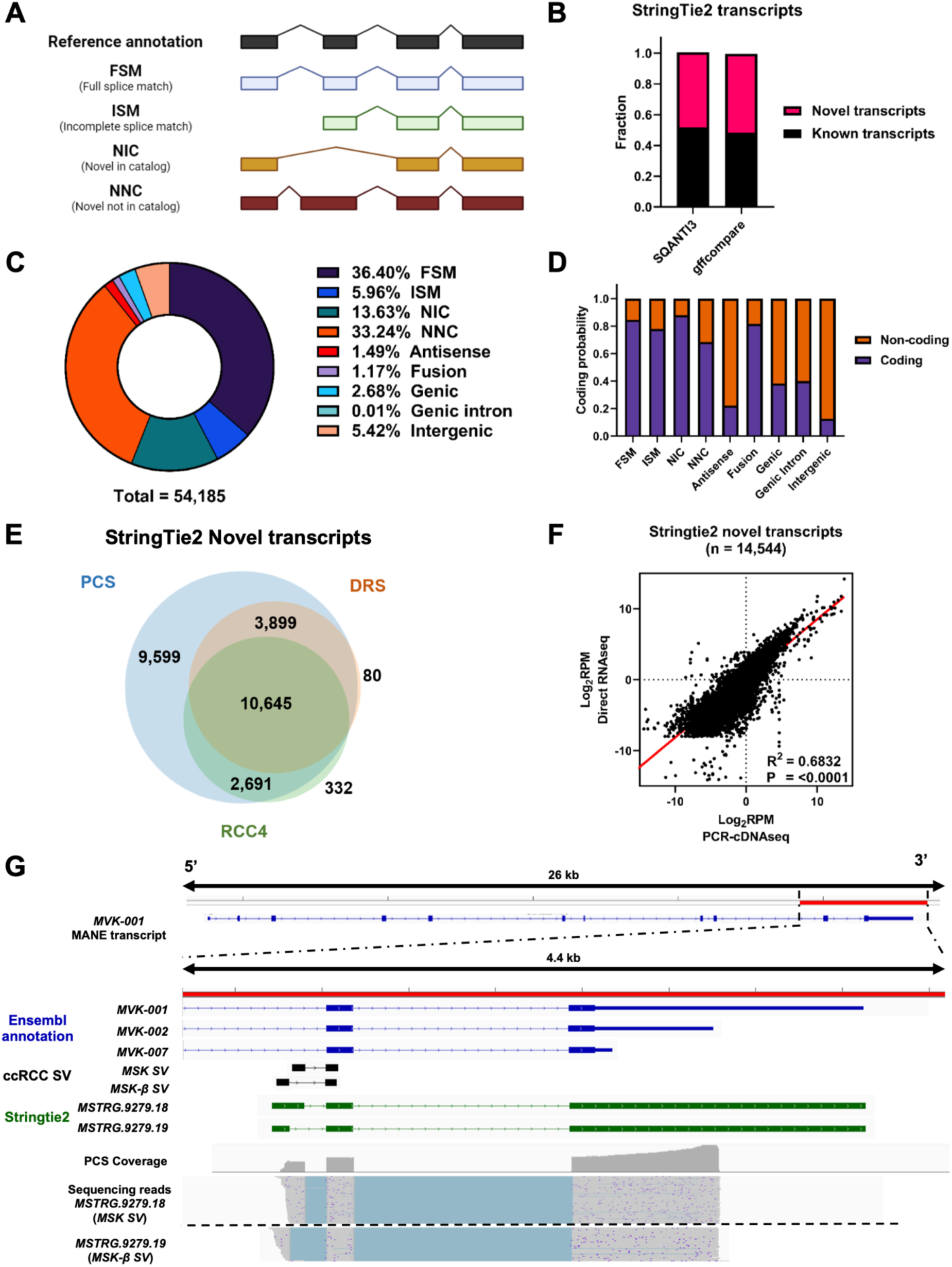
Long-read RNA sequencing enables the discovery of full-length novel transcripts. **A**. Graphical representation of the major SQANTI3 isoform categories (antisense, genic intron, genic genomic and intergenic not shown here). **B.** Bar chart showing the proportion of Novel and Known transcripts in Stringtie2 assembly as curated by SQANTI3 and gffcompare **C** Pie chart depicting the distribution of SQANTI3 isoform categories amongst Stringtie2 assembled transcripts (n = 54,185). **D.** Bar chart showing the proportion of coding and non-coding Stringtie2 assembled transcripts by SQANTI3 isoform categories. **E.** Venn diagram showing the number of overlapping mapped StringTie2 novel transcripts between PCS and DRS of ccRCC tumour samples, and DRS of ccRCC cell line RCC4. **F.** Correlation between transcripts expression levels (Log_10_ Reads per million (RPM)) of all StrinTie2 novel transcripts mapped by both PCS and DRS (n = 14,544). Diagonal line represents the line of best fit. R^2^ value was computed to measure goodness-of-fit and p value was generated from F-test, with p<0.05 considered statistically significant. **G** IGV visualisation of *MVK* reference annotations (blue), ccRCC specific *MVK* splice junctions (black), Stringtie2 assembled novel transcripts (green), PCS coverage track (grey) illustrating the depth of sequence coverage across the region of interest (red bar, hg38 chr12:109,594,200 – 109,598,600) and PCS sequencing reads aligned to the reference genome in the region of interest.

Remarkably, both SQANTI3 and gffcompare classifications revealed that novel transcripts constitute more than 50% of the assembled transcripts from the nephrectomy specimens (**Fig. 3B**). For SQANTI3 classification, the most prominent class of assembled transcripts were FSM (36.5%, n = 19,722, **Fig. 3C**). Interestingly, within the FSM isoforms, 15.3% exhibited an alternative 3’ end (n = 3,010), 11.0% contain an alternative 5’ end (n = 2,170) and 4.9% of FSM transcripts display alternative 3’ and 5’ ends (n = 962) when compared to the reference annotation (**Additional file 1: Table S7**). Under a broader classification criterion, these isoforms could be considered as putative novel transcripts. Importantly, analysis by SQANTI3 also indicated that the large proportion of novel transcripts may possess coding potential (**Fig. 3D**).

Similarly, gffcompare analysis revealed that the predominant class of StringTie2 assembled transcripts was ‘j’ (Novel, multi-exon gene with at least 1 matched exon junction) (40.58%, n = 21,635), followed by ‘=’ (Known, complete intron chain match) (28.32%, n = 15,099) (**Additional file 1: Table S8**). Applying the alternative long-read RNAseq transcript assembler

FLAIR on PCS reads, gffcompare characterisation reaffirmed that the majority of assembled transcripts are novel isoforms (**Additional file 1: Table S8**). Detailed characterisations of novel Stringtie2 assembled transcripts by SQANTI3 and gffcompare can be found in **Additional File1: Table S9**.

Next, we asked whether the novel assembled transcripts can also be detected by DRS of nephrectomy samples and, importantly, ccRCC tumour cells *in vitro*. The latter could indicate the cancer cell-intrinsic origin of these transcripts. To address this, we performed DRS analysis of the *VHL*-negative, ccRCC cell line RCC4 under both untreated and IFNγ and TNF treated conditions using DRS. The cytokine treatment conditions aimed to simulate in part the transcriptomic response of tumour cells to immune cells. Workflow and sequencing statistics can be found in **Fig. S7**, and DEG analysis data can be found in **Additional File1: Table S10**. Out of the 26,834 novel isoforms that were mapped by PCS of nephrectomy samples, 14,544 were also mapped in at least one DRS of the nephrectomy samples, and 13,336 were also detected in the DRS of RCC4 samples (**Fig. 3E**). Furthermore, expression levels of these novel isoforms exhibited strong concordance between PCS and DRS of nephrectomy samples (R^2^ = 0.6832, P = <0.0001) (**Fig. 3F**). Despite starting with a reference based on our PCS dataset, a small number of StringTie2 transcripts were mapped exclusively in DRS of nephrectomy samples (n = 80) and RCC4 (n = 332). This may be due to the assignment of multi-mapping, ambiguous sequencing reads by the sequence alignment program (minimap2). These results reveal a plethora of previously uncharacterised and unmapped transcripts within the ccRCC transcriptome. Despite differences in sequencing depth, novel transcripts from PCS could also be mapped by DRS, with a substantial proportion of these novel transcripts also detected in ccRCC cells *in vitro*.

### Long-read RNA sequencing reveals the full exonic structure of ccRCC splice variants

Taking advantage of the ability of long-read sequencing to reveal whole transcript exonic structures, we next examined recently reported novel, non-reference annotated SVs specific to ccRCC tumours, which were supported by proteomics data and associated with clinical outcomes [44]. We found sequence read evidence for all 16 reported SVs (15/16 for PCS of nephrectomies, 9/16 for DRS of nephrectomies and 13/16 for DRS of RCC4). Moreover, PCS StringTie2 assembled transcripts spanning 11 of the unannotated SVs (**Additional file 1: Table S11**). For example, the StringTie2 assembled transcripts *MSTRG.9279.18* and *MSTRG.9269.19* accurately replicated two ccRCC-specific SVs from *MVK* (**Fig. 3G**). This was supported by reference genome aligned reads from all three long-read RNAseq datasets (**Fig. S8A**). In addition, sequencing results showed that these ccRCC splice variants adopt the 3’UTR structure of *MVK-002* (ENST00000392727) instead of the longer 3’UTR from canonical MANE transcript *MVK-001* (ENST00000228510). Another example was *HPCAL1*, where two StringTie2 assembled transcripts (*MSTRG.20400.11* and *MSTRG.20400.12*) were found to span the ccRCC-specific SV (**Fig. S8B**). The two isoforms exhibit variation in the exon 3 usage, where evidence of exon 3 retention can be found in PCS as well as RCC4 sequencing results. Overall, 3 additional SVs (*SYNPO*, *EGFR* and *FAM107B*) were found to be encompassed by 2 StringTie2 assembled transcripts (**Additional file 1: Table S11**). Collectively, using recently reported ccRCC-related SVs [44], we demonstrated the ability of long-read sequencing to reveal transcriptomic co-dependencies, in this case the co-occurrence of specific SVs with specific UTRs, providing unparalleled insight into novel features of ccRCC.

### Discovery of a novel soluble PD-L1 isoform expressed by ccRCC tumour cells

Having identified that a reduction in the immune infiltrate of ccRCC tumours was linked to disease recurrence and that the ccRCC transcriptome includes a high number of previously uncharacterised novel transcript isoforms we focused on transcripts of immune checkpoint proteins. Here, we focused on programmed death-ligand 1 (PD-L1). Whilst most studies on PD-L1 have focused on the membrane-bound isoform (*mPD-L1*), recent attention has been drawn to a soluble PD-L1 isoform (*sPD-L1*) lacking exon 5, 6, and 7. sPD-L1 is currently unannotated in the Ensembl gene annotation, but it has been described in the NCBI GenBank database (NM_001314029). An Ensembl annotated transcript (ENST00000474218) partially overlaps with the 3’UTR of the GenBank sPD-L1 transcript, serving as a proxy for mapping *sPD-L1*. [45]. Upon closer inspection to the exon 4 and 3’UTR region of *sPD-L1* (chr5: 5,462,800 – 5,463,400), reference genome reads coverage and StringTie2 supported two distinct isoforms with varying 3’UTR lengths (**Fig. 4A**). While the shorter *sPD-L1* represents the GenBank transcript, the alternative *sPD-L1* includes a 3’UTR more than twice the length of GenBank annotation (61nt vs 154nt) (**Fig. 4A**). StringTie2 assembly revealed this elongated 3’UTR structure, with supporting evidence stemming from reference genome aligned reads of PCS and DRS data from ccRCC tissues, as well as DRS data from RCC4 cells (**Fig. S9**).

**Figure 4:**
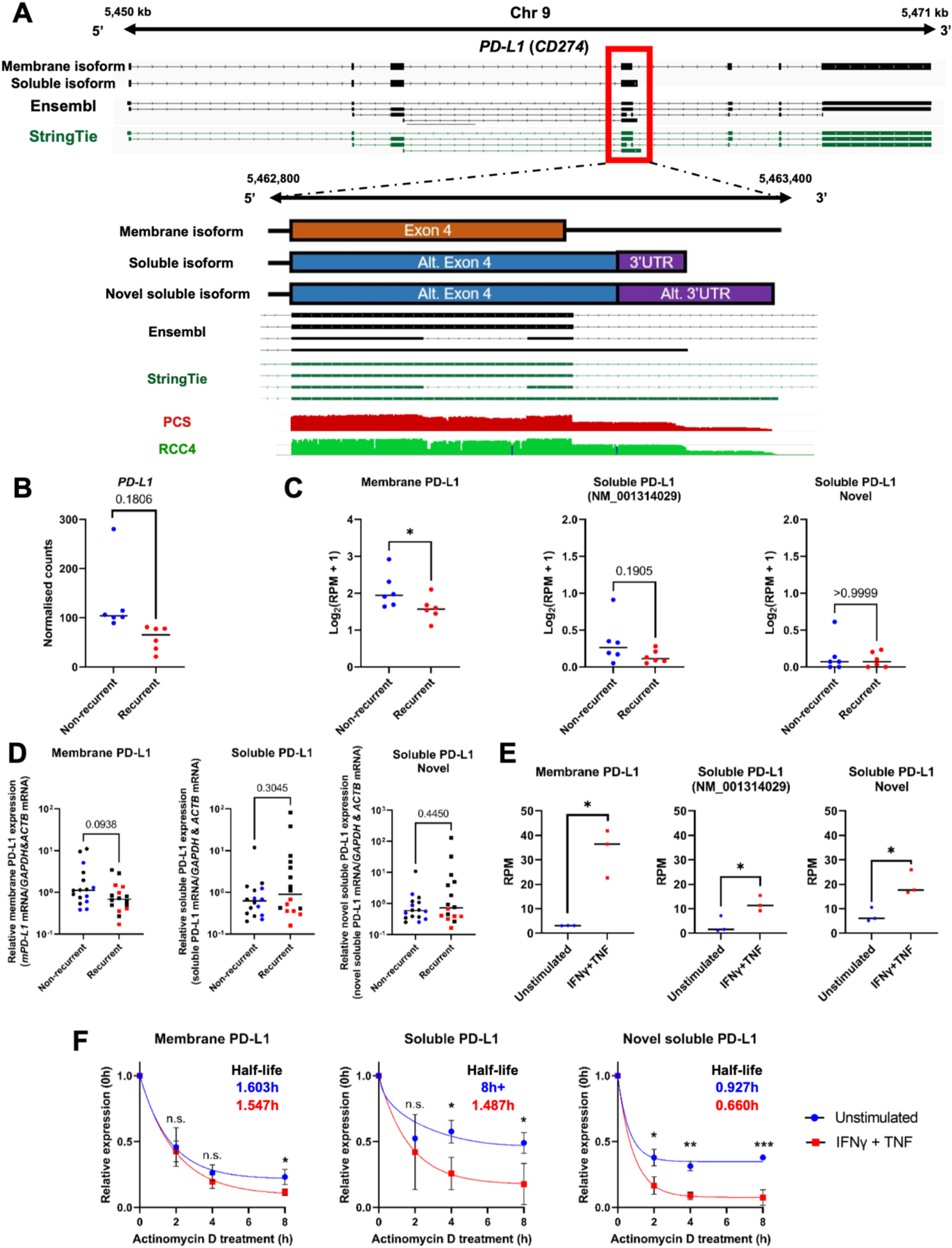
Discovery of a novel *sPD-L1* isoform expressed by ccRCC tumour cells. **A.** IGV visualisation of reference annotation of *mPD-L1* isoform (black, ENST00000381577), *sPD-L1* (black, NM_001314029) and Stringtie reference annotation (green) (Top tracks); Graphical representation of membrane, soluble and novel soluble *PD-L1* exon 4; Ensembl (black) and Stringtie reference annotations (green) and IGV coverage tracks for PCS of ccRCC tumours (red) and DRS of RCC4 (green). **B.** Grouped dot plot showing reference DESeq2 normalised *PD-L1* expression in non-recurrent (blue) and recurrent (red) tumours’ PCS data. DESeq2 p_adj_ value is shown in graph. Centre line represents median for each group. **C.** Grouped dot plots showing normalised *mPD-L1*, *sPD-L1* and novel *sPD-L1* expression (log_2_(RPM+1)) in non-recurrent (blue) and recurrent (red) tumours’ PCS data. **D.** Grouped dot plots showing *mPD-L1*, *sPD-L1* (all isoforms) and novel *sPD-L1* mRNA levels measured by qRT-PCR in recurrent and non-recurrent tumours from sequenced cohort (blue and red, n = 12) and validation cohort (black, n = 20) relative to average mRNA levels in non-recurrent tumours. **E.** Grouped dot plot normalised *mPD-L1*, *sPD-L1* and novel *sPD-L1* expression in unstimulated (blue) and IFNγ+TNF treated (red) RCC4 cells. For **C – E**, two-tailed Mann-Whitney U tests were used with p ≤ 0.05 considered significant. * = p < 0.05. Centre line represents median for each group. **F.** mRNA decay curves for *mPD-L1*, *sPD-L1* and novel *sPD-L1* in unstimulated (blue) and IFNγ + TNF treated (red) RCC4 cells. Half-lives of isoforms are indicated in graph (blue for unstimulated, red for IFNγ+TNF treated RCC4). Comparisons were made using unpaired student’s T test with p ≤ 0.05 considered significant. n.s. = not significant, * = p < 0.05, ** = p < 0.01, *** = p < 0.001.

Upon evaluating the expression dynamics of *PD-L1* in ccRCC tumours, no significant disparity in gene-level expression was found between recurrent and non-recurrent nephrectomy samples (**Fig. 4B**). However, at the isoform level, whilst *mPD-L1* was suppressed in the recurrent samples, both *sPD-L1* isoforms (NM_001314029 and novel sPD-L1) showed no significant differences (**Fig. 4C**). Subsequent expression validation via qRT-PCR with the sequenced and independent validation cohort displayed the same pattern of results, where *mPD-L1* displayed a borderline non-significant (p=0.09) down regulation, while *sPD-L1* isoforms remained unchanged (**Fig. 4D, S10**).

As all *PD-L1* transcripts, including the novel *sPD-L1* isoform, were detected in RCC4 cells by DRS, we sought to further explore their regulation in cancer cells. The expression of all *PD-L1* isoforms increased in response by IFNγ+TNF treatment (**Fig. 4E**). However, expression levels of *mPD-L1* were profoundly more responsive to cytokine treatment than the soluble isoforms (approximately 30-fold induction of *mPD-L1* as opposed to 3 – 10-fold induction of *sPD-L1* isoforms). Intriguingly, mRNA stability assays revealed that cytokine treatment significantly reduced the stability of *sPD-L1* but not *mPD-L1* (**Fig. 4F**). Furthermore, the novel *sPD-L1* isoform exhibited a notably lower stability than the total *sPD-L1* isoforms. Taken together, our findings revealed the existence of an up-to-now uncharacterised *sPD-L1* isoform with a longer 3’UTR and low stability, and key differences in the regulation of membrane and soluble *PD-L1* isoforms in ccRCC tumours and in response to inflammatory cytokines *in vitro*.

### Discovery of novel genes associated with ccRCC recurrence

In addition to the characterisation of novel isoforms within known genes, long-read RNAseq also enables the discovery of novel genes that are absent from the reference gene annotation. Using the PCS StringTie2 assembly, we identified 1,350 novel genes (curated by SQANTI3) that were mapped in the PCS dataset. The majority of these genes were classified as either intergenic (59.76%) or antisense (39.86%) transcripts (**Fig. 5A**). Most of these novel genes have a single isoform, with the majority being non-coding, multi-exon isoforms featuring canonical splice sites (**Fig. 5B**, **S11A – C**). The expression levels of these novel genes are similar to those of reference annotated genes, with the coding novel genes demonstrating higher expression levels than non-coding novel genes (**Fig. S11D**). Importantly, of the 1,350 novel genes that were mapped by PCS, 982 (72.7%) were also detected in the DRS data of tumour nephrectomies, and 414 (30.7%) novel genes were also mapped in the DRS data of RCC4 cells (**Fig. 5C**). This suggests that a large number of novel genes might be expressed in ccRCC tumour cells.

**Figure 5:**
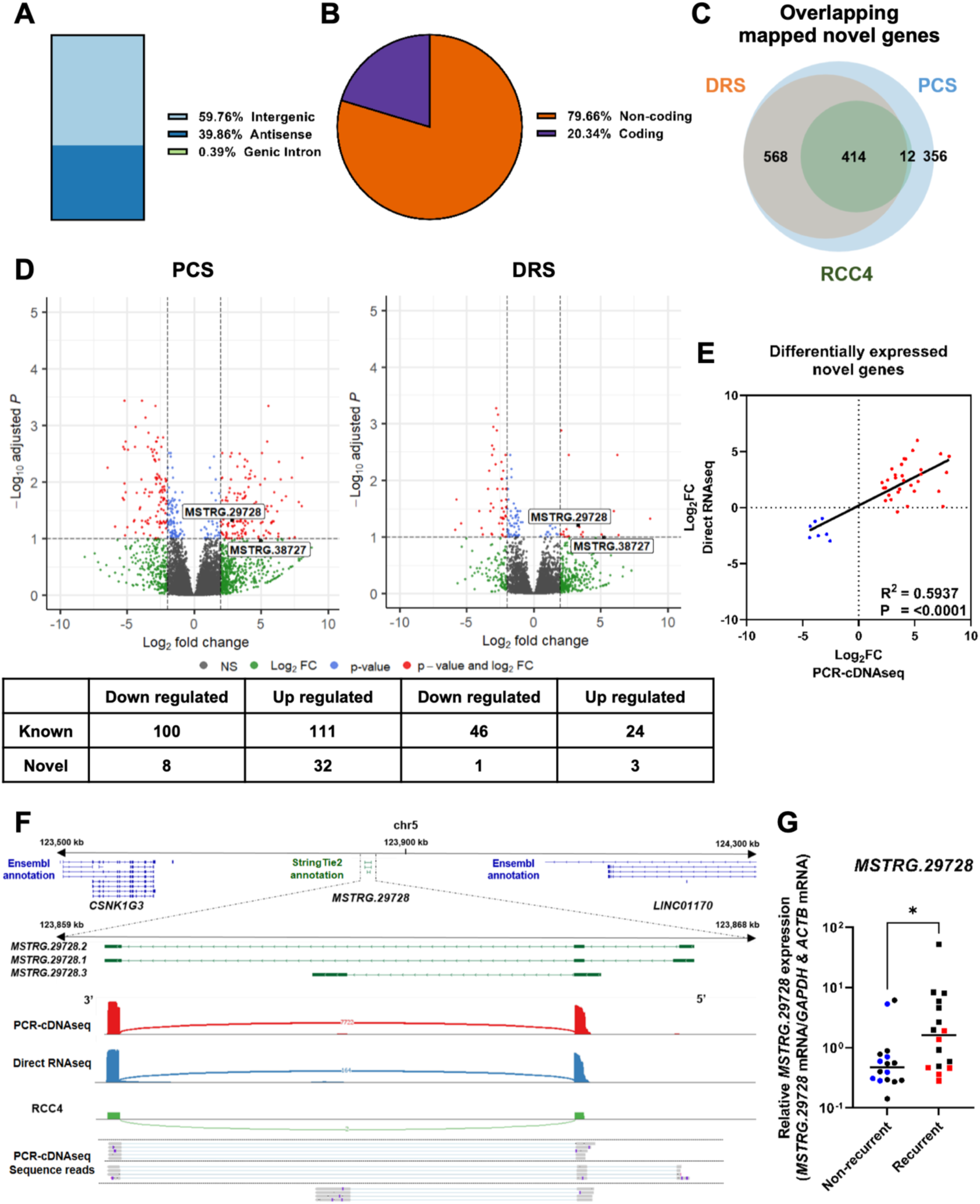
Discovery of ccRCC recurrence associated novel genes by long-read RNAseq. **A**. Bar chart showing the isoform classifications of Stringtie assembled transcripts from novel genes as classified by SQANTI3. **B.** Pie chart illustrating the proportion of coding and non-coding Stringtie assembled transcripts from novel genes as classified by SQANTI3. **C.** Venn diagram showing the number of overlapping mapped novel genes between PCS and DRS of ccRCC tumour samples, and DRS of RCC4. **D.** Volcano plots showing differentially expressed genes (red) between recurrent and non-recurrent tumours from PCS and DRS data using StringTie2 assembled reference. Number of differentially expressed novel and known genes are shown in table below plots. Names of novel genes that were validated by qPCR with validation cohort are shown on plots. **E.** Correlation between log_2_FoldChange of differentially expressed novel genes identified by either or both PCS and DRS between recurrent vs non-recurrent tumours (n = 40). **F.** IGV visualisation of *MSTRG.29728* isoforms StringTie2 reference annotation (green) and the closest neighbouring genes (*LINC01170* and *CSNK1G3*) in the Ensembl reference annotation (Ensembl release 105) at chr5:123,500,000-124,300,000 (Top track); Sashimi plot showing abundance of reference genome aligned reads and splicing patterns along *MSTRG.29728* (chr5:123,859,000-123,868,000) for PCS (red) and DRS (blue) of ccRCC tumour samples, and DRS of RCC4 (green); Representative PCS sequencing reads (grey) aligned to the reference genome in the region of interest. **G.** *MSTRG.29728* mRNA levels measured by qRT-PCR in recurrent and non-recurrent tumours from sequenced cohort (blue and red, n = 12) and validation cohort (black, n = 20), relative to average mRNA levels in non-recurrent tumours. mRNA levels were normalised to *GAPDH* and *ACTB*. Two-tailed Mann-Whitney U test was used with p ≤ 0.05 considered significant. * = p < 0.05. Centre line represents median for each group.

Next, we performed DEG analysis with the PCS StringTie2 assembly and identified a set of significantly differentially expressed (|log_2_FoldChange| ≥ 2, p_adj_ ≤ 0.1) novel genes (n = 40 for PCS, n = 4 for DRS) between recurrent and non-recurrent samples (**Fig. 5D, S11E, Additional File1: Tables S12-15**). The directionality of gene expression for these differentially expressed novel genes demonstrated strong concordance between PCS and DRS (**Fig. 5E**). 13 differentially expressed novel genes were also identified between untreated and IFNγ+TNF treated RCC4 cells (**Fig. S11F**).

To further validate our sequencing findings, we sought to experimentally measure the levels in recurrent and non-recurrent ccRCC nephrectomies of two novel genes: *MSTRG.29728* and *MSTRG.38727* using qRTPCR. *MSTRG.29728*, a StringTie2 assembled gene is located on chromosome 5, with its nearest reference annotated genes (5’: *CSNK1G3*, 3’: *LINC01170*) situated more than 300 kb away (**Fig. 5F**). Notably, the presence of this novel gene was also supported by reference genome aligned reads from the DRS RCC4 datasets. *MSTRG.29728* was significantly up-regulated in recurrent ccRCC tumours in both PCS and DRS datasets.

This up-regulation was confirmed by qRT-PCR in both sequenced and independent validation cohorts (**Fig. 5G**). The second tested novel gene, *MSTRG.38727,* is located on chromosome X with read coverage from PCS and DRS of nephrectomy specimens, albeit absent in DRS data from RCC4 (**Fig. S12A – C**). PCS sequencing results showed that *MSTRG.38727* expression was highly elevated in 3 of the 6 recurrent ccRCC tumours (**Fig. S12D**). This was corroborated through qRT-PCR validation within the sequenced cohort, but was not validated in the independent validation cohort (**Fig. S12E**).

Overall, long-read sequencing revealed a high number of candidate novel genes present in ccRCC transcriptomes. Further testing for two such genes by orthogonal methods and in independent patient cohorts provide further support for their existence and, critically, identified *MSTRG.29728* as a novel non-coding RNA gene associated with ccRCC recurrence in both study cohorts. We provisionally term *MSTRG.29728* as *RECART*, for *Renal Carcinoma Recurrence associated Transcript*.

## Discussion

Long-read sequencing technologies represent a new era in cancer genomics and RNA medicine [46, 47]. Long-read RNA sequencing in particular can reveal how co-dependent of co– and post-transcriptional features contribute to transcriptome plasticity. In the context of cancer, this plasticity, also referred to as transcriptome instability, is thought to underpin oncogenesis and disease progression but also the interaction between cancer cells and the immune system immunogenicity [17, 18]. Driven by these observations, we used DRS and PCS to explore transcriptomes of primary ccRCC tumours.

From a methodological point of view, the distinguishing features of our study are (i) the direct comparison between DRS and PCS of clinical samples, (ii) the successful sequencing of archival fresh frozen tissue samples, and (iii) the use of total RNA as starting material for DRS and PCS library preparation. In reference to the latter point, compared to the pg-ng range of total RNA input requirement for short-read RNA sequencing library preparation, previous studies using ONT DRS have typically used 50 – 500 ng of poly(A) enriched RNA, which is hugely demanding for clinical samples [48]. Here we used 2 μg and 200 ng total RNA for DRS and PCS from tissues, respectively without poly(A) enrichment. We reasoned that since the first step of DRS and PCS library generation involves binding an adapter primer containing a poly-d(T) sequence, a prior step for the enrichment of poly(A) RNAs should not be necessary. Indeed, a recent report has demonstrated that poly(A) selection can also introduce a potential bias towards mRNAs with longer poly(A) tails [49]. PCS achieved a higher depth and, consequently, detected a higher number of transcripts and genes in all tested samples, and a higher number of DEGs in primary tumours of patients who experienced recurrence than DRS. We note that we did not multiplex samples for PCS. Importantly however, DRS does not include a PCR amplification step. This provides further confidence in the overlapping gene sets between the two methods and can also explain the small number of genes and transcripts or DEGs detected only by DRS. With regards to read-length, both methods produced long reads. On average, raw reads generated by PCS were longer than DRS reads likely because of the fact that raw reads from PCS have additional ligated reverse transcription, PCR amplification primer and unique molecular identifier. Once aligned to the reference genome, both methods achieved similar read lengths, although PCS achieved a higher percentage of full-length transcripts likely due to the size selection step following PCR [50]. However, it is important to note that the successful use of DRS in archival clinical samples presented here opens the route for additional analyses including estimation of polyA length per transcript and post-transcriptional RNA modifications [51]. These were not performed here because the focus of the study was on novel transcript and gene identification and comparisons and overlaps between DRS and PCS.

The primary biological objective of our study was to use DRS and PCS to explore ccRCC recurrence-associated transcriptome features including previously uncharacterised genes and transcripts. Our differential expression and deconvolution analyses identified a loss of immune infiltrate and specifically CD8^+^ T cells as a key feature of primary tumours that go on to relapse after surgery. This is reported by others [52, 53] providing a biological validation of our findings. Interestingly, and despite the low numbers of samples tested in our sequencing cohort we were able to see similar expression patterns for NOS3 and CCL5, two previously reported recurrence markers [14]. In addition, we found that down-regulation of *TOX* was linked to recurrence (note that *TOX* levels were not measured in the study that identified loss of *NOS3* and and *CCL5* as a recurrence marker). TOX is a transcription factor initially identified as a marker of exhausted CD8^+^ T cells [54, 55], although it has also been shown that it is expressed in all human effector CD8^+^ T cells [56]. This might in part explain why only *TOX* but not *PD1* was associated with recurrence in our validation cohort. Importantly, in addition to these already known genes, our study identified upregulation of a novel gene, *MSTRG.29728* or *RECART*, as an event linked to disease relapse. This predicted lincRNA was detected by both PCS and DRS and it is also expressed in RCC4 cells, suggesting a potential cancer cell-intrinsic origin and setting the foundation for future studies on its function in ccRCC.

A unique strength of long-read sequencing is the ability to determine transcript co-dependencies, for example preferential use of specific UTRs by specific SVs, which can suggest tissue– or disease-specific co-transcriptional processing mechanisms. Indeed, we demonstrated this for a selection of recently identified ccRCC-associated SVs [44], including *MVK* and *HPCAL1*. Focusing these analyses on immune checkpoints led to the discovery of a novel *sPD-L1* transcript with a longer 3’UTR than the currently annotated *sPD-L1*. This means that the novel *sPD-L1* is likely to be controlled by additional post-transcriptional mechanisms, including microRNA-mediated silencing or regulation by RNA-binding proteins. Of note, regulation through the 3’UTR is a major determinant of mPD-L1 expression [57, 58]. Indeed, the novel *sPD-L1* transcript demonstrates lower stability than the other *PD-L1* transcripts under homeostatic or inflammatory conditions *in vitro*. Interestingly, we found that there is a trend for down-regulation for tumour *mPD-L1* but no differences in *sPD-L1* in patients that experience recurrence. This is consistent with the observed loss of CD8^+^ T cells from these tumours and the enhanced responsiveness of *mPD-L1* to IFNγ and TNF observed *in vitro*. Clinically, this is important as PD-1/PD-L1-targeted checkpoint inhibitors are currently being explored in the adjuvant setting, with inconsistent results reported to date [59–62] and expression of sPD-L1 has been linked with ccRCC prognosis and immunotherapy treatment outcome [63, 64]. Future studies will have to explore the relative contributions of the different *PD-L1* transcripts, including the novel one reported here, to tumour immune evasion and response to immunotherapies.

Even though we analysed a sequencing and an independent validation cohort, the relatively low total number of study participants (n = 32) is a limitation of our study that should be considered. As such future studies will need to further validate our results also considering the ccRCC genetic alteration hierarchy and tumour heterogeneity.

## Conclusion

Our results provide a fundamental redefinition of the ccRCC transcriptome including thousands of novel transcript isoforms and hundreds of novel genes detected by both DRS and PCS in primary ccRCC tumours but also *in vitro* in ccRCC cell lines. In agreement, with others we show that immune cell depletion from the primary tumour is associated with disease relapse. We report loss of the exhausted CD8^+^ T cell marker *TOX* and upregulation of *RECART*, a novel lincRNA, as novel indicators of relapse. Strikingly, we also discover a novel *sPD-L1* isoform with differential stability and we show that only *mPDL1* might be deregulated in primary tumours of patients who experience relapse. These findings demonstrate that long-read sequencing has the potential to lead to a radical revision of our understanding of the plasticity and instability of cancer transcriptomes.

## Availability of data and materials

Material: The sequencing results are available as raw reads and the gene counts tables are available at gene expression omnibus as below: PCS GSE242204, DRS GSE241932, RCC4 DRS: GSE242084.

Code: The workflows for reads alignment are available at https://github.com/joshuacylee/DRS and https://github.com/joshuacylee/PCR-cDNAseq. Workflow for differential gene expression identification is available at (https://github.com/joshuacylee/DESeq2).

## Data Availability

Material: The sequencing results are available as raw reads and the gene counts tables are available at gene expression omnibus as below: PCS GSE242204, DRS GSE241932, RCC4 DRS: GSE242084.
Code: The workflows for reads alignment are available at https://github.com/joshuacylee/DRS and https://github.com/joshuacylee/PCR-cDNAseq.
Workflow for differential gene expression identification is available at (https://github.com/joshuacylee/DESeq2).

## Acknowledgements

We are indebted to the study participants and their families for contributing to medical research. We thank Aino Järvelin for support with bioinformatics analyses of long-read sequencing data.

## Contributions

JL: experimental design; data generation, data analysis, figure design, and manuscript writing. EAS: experimental design; data generation. JB: clinical sample and data collection and maintenance. REB: clinical sample and data collection. DJT: study conception; study design. NSV: clinical sample and data collection; study design; data interpretation. DL: study conception; study design; data interpretation; study supervision; manuscript writing; All authors read and approved the final manuscript.

## Ethics declarations

Ethics approval and consent to participate

This study was approved regional ethics committee approval: Yorkshire & The Humber – Leeds East Research Ethics Committee, reference 15/YH/0080. The research conforms with the principles of the Declaration of Helsinki. All patients gave written informed consent for their participation in this study.

## Consent for publication

Not applicable.

## Supplementary information

**Additional File 1: Supplementary Tables Table S1:** Clinical characteristics of ccRCC patients from sequenced cohort (n = 12) and validation cohort (n = 20). **Table S2:** List of primers. **Table S3:** Sequencing output statistics. **Table S4:** PCS recurrent vs non-recurrent ccRCC differential gene expression analysis. **Table S5:** DRS recurrent vs non-recurrent ccRCC differential gene expression analysis. **Table S6:** Gffcompare class codes and SQANTI3 categories equivalence. **Table S7:** SQANTI3 statistics on StringTie2 assembly. **Table S8:** Gffcompare statistics on StringTie2 and FLAIR assembly. **Table S9:** SQANTI and Gffcompare characterisation of Stringtie2 transcripts. **Table S10:** RCC4 untreated vs IFNγ+TNF treated DRS differential gene expression analysis. **Table S11:** Identification of ccRCC specific splice junctions. **Table S12:** PCS recurrent vs non-recurrent ccRCC Stringtie2 differential gene expression analysis.

## Additional File 2: Supplementary Figures

Figure S1: DRS and PCS of ccRCC nephrectomy samples. **Figure S2:** Correlation between PCS and DRS data. **Figure S3:** Principal Component Analysis (PCA) plots on ccRCC tumours gene expression data. **Figure S4:** Recurrence of ccRCC is associated with lower tumour immune infiltration. **Figure S5:** Validation of sequencing results via qRT-PCR. **Figure S6:** Graphical representation of GffCompare transcript class codes. **Figure S7:** Sequencing statistics of IFNγ + TNF treated and untreated RCC4 cells. **Figure S8:** Visualisation of long-read RNAseq reads mapping to ccRCC-specific splice junctions. **Figure S9:** Sequencing reads aligned to *PD-L1* exon 4. **Figure S10:** Validation of *PD-L1* sequencing results and novel *sPD-L1* isoform via qRT-PCR. **Figure S11:** Characterisation of novel genes. **Figure S12:** Visualisation and expression validation of novel gene *MSTRG.38727*.

