## Supplementary Figures for "Long-read RNA sequencing redefines the clear cell renal cell carcinoma transcriptome and reveals novel genes and transcripts associated with disease recurrence and immune evasion"

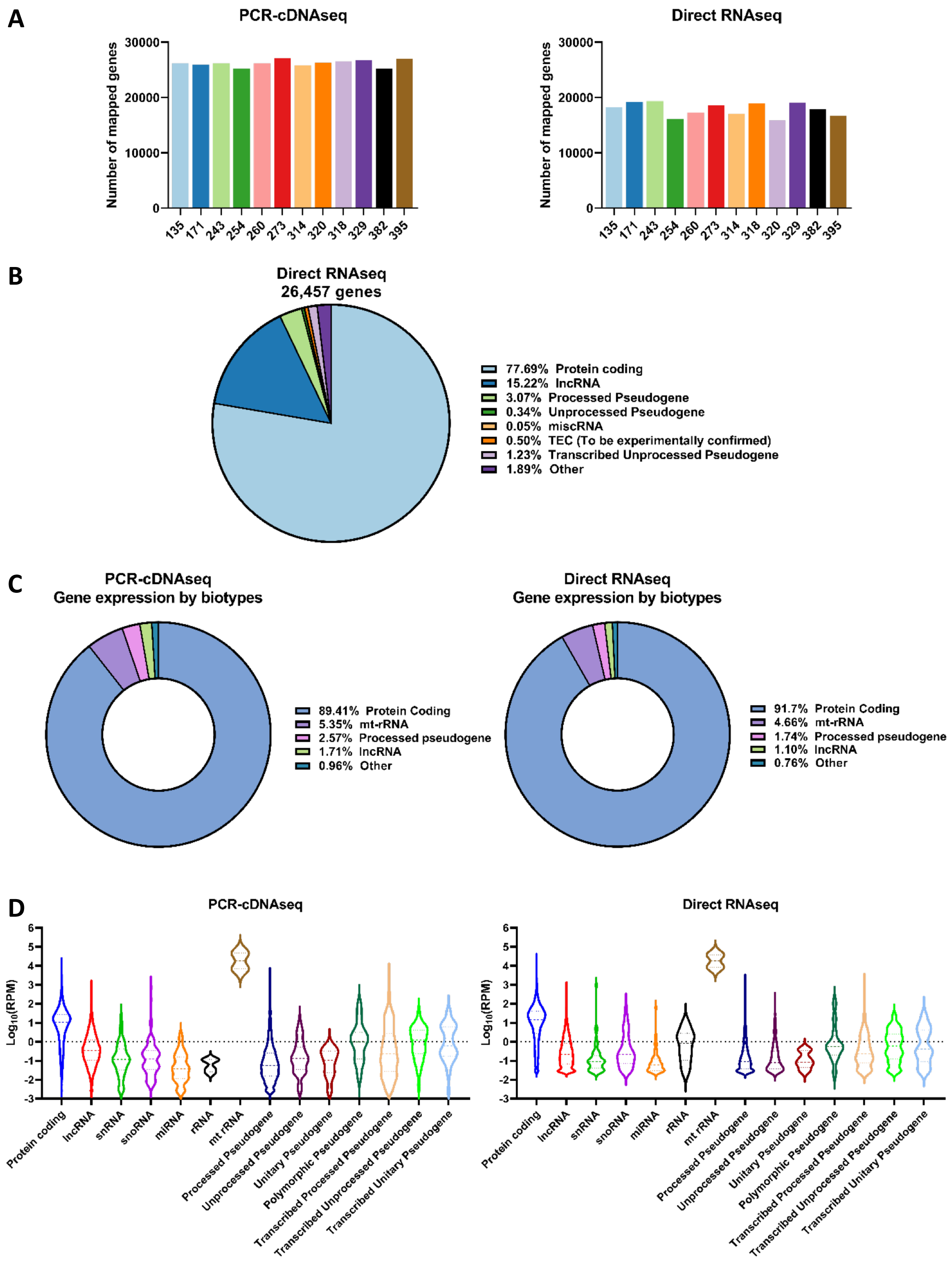


**Figure S1: DRS and PCS of ccRCC nephrectomy samples. A.** Bar charts showing the number of genes mapped by PCS and DRS from each ccRCC tumour sample. **B.** Pie chart depicting the proportions of gene biotypes of all mapped genes from reference genome

(Ensembl release 105, GRCh38) by DRS of sequenced tumour samples. **C.** Pie charts depicting the average proportions of RNA biotypes of mapped genes by expression levels from PCS and DRS data. **D.** Violin plots depicting the distribution of gene expression levels ($Log_{10}$RPM) of mapped genes by biotypes from PCS and DRS, with first and third quartiles and median shown as horizontal line within each plot.

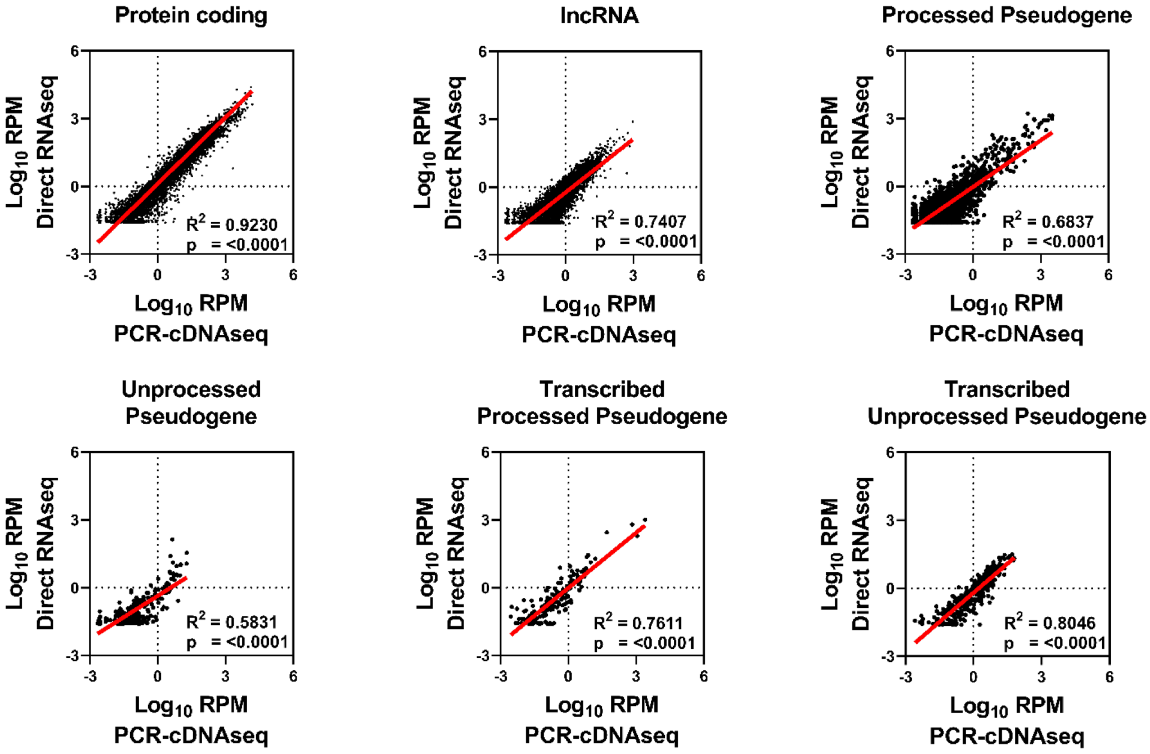


**Figure S2: Correlation between PCS and DRS data.** Correlation between gene expression levels ($Log_{10}$RPM) of Protein coding, lncRNA, Processed Pseudogene, Unprocessed Pseudogene, Transcribed Processed Pseudogenes and Transcribed Unprocessed Pseudogenes mapped by PCS and DRS of ccRCC tumour samples. Throughout, diagonal lines represent the line of best fit. $R^2$ values were computed to measure goodness-of-fit and p values were generated from F-test, with $p<0.05$ considered statistically significant.

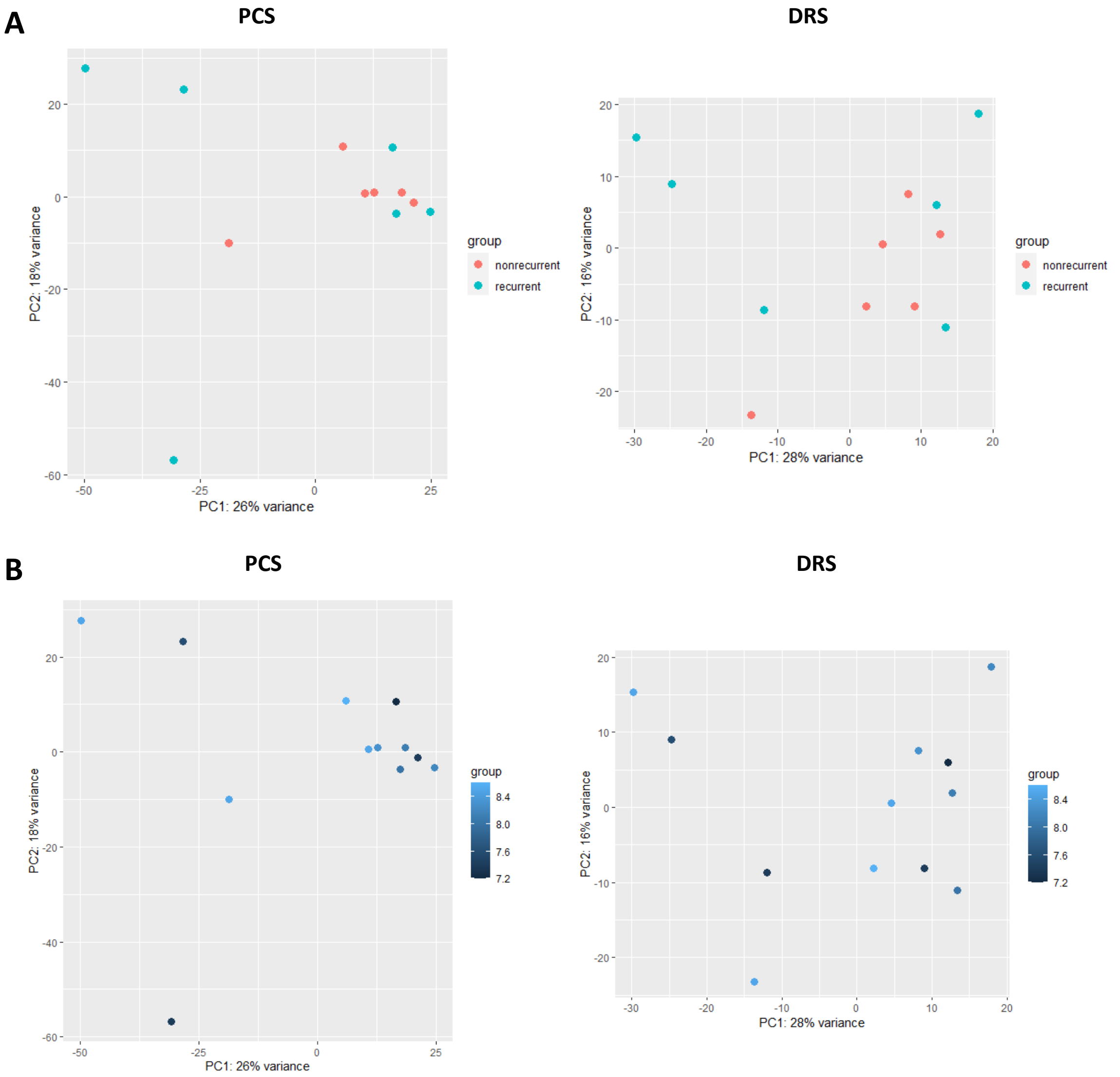


**Figure S3: Principal Component Analysis (PCA) plots on ccRCC tumours gene expression data. A.** DESeq2 generated PCA plots using PCS and DRS expression data showing PCA of recurrent vs non-recurrent ccRCC groups. **B.** PCA plots using PCS and DRS expression data showing PCA of samples with respective RIN numbers.

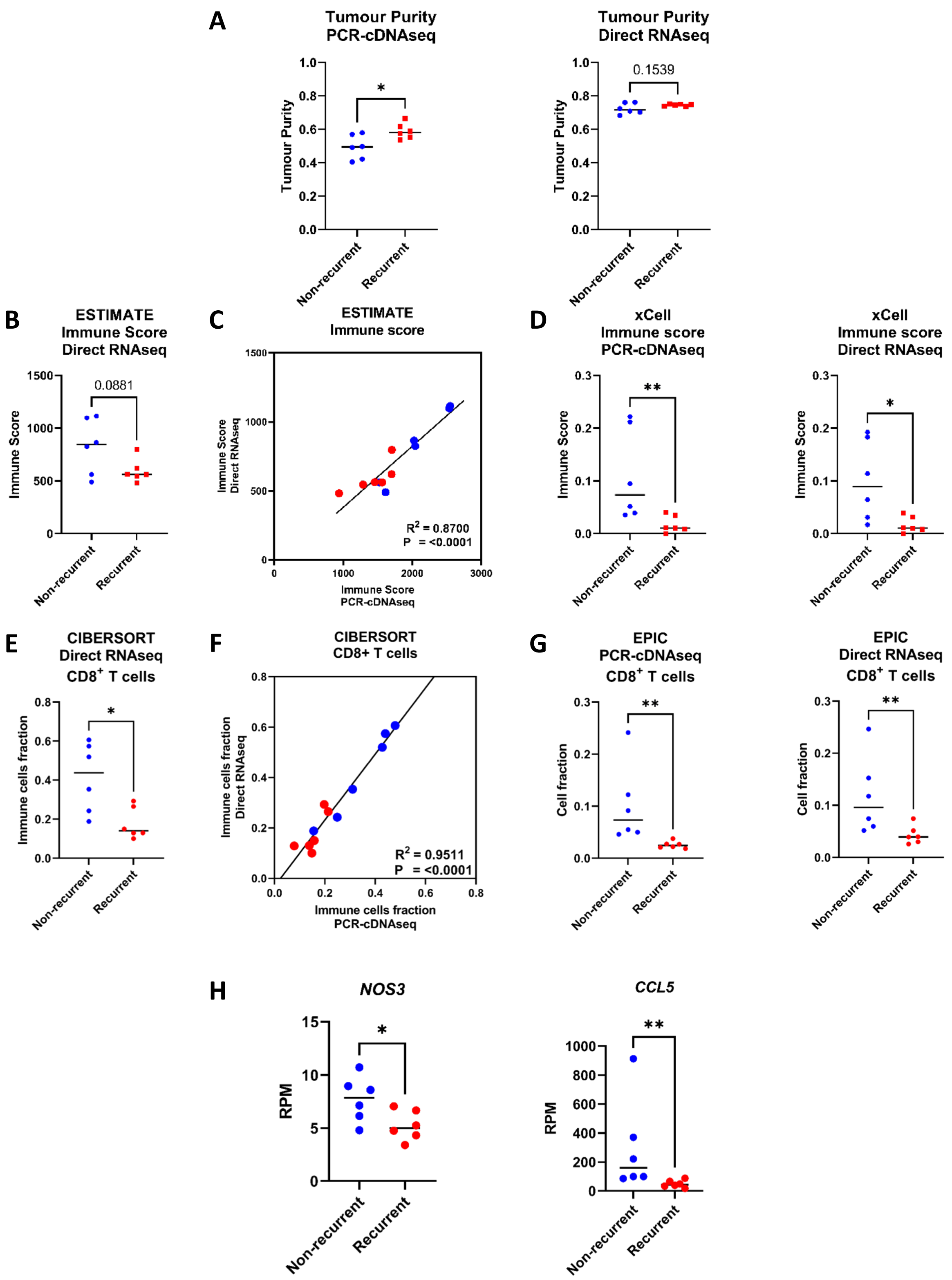


**Figure S4: Recurrence of ccRCC is associated with lower tumour immune infiltration.** **A.** Grouped dot plot showing estimated tumour purity of non-recurrent (blue) and recurrent (red) ccRCC tumours by the ESTIMATE algorithm using gene expression data from PCS and DRS. **B.** Grouped dot plot showing estimated immune score of non-recurrent (blue) and recurrent (red) ccRCC tumour by the ESTIMATE algorithm, using DRS gene expression data. **C.** Correlation between ESTIMATE immune scores of non-recurrent (blue) and recurrent (red) ccRCC tumours, generated by PCS and DRS gene expression data. **D.** Grouped dot plot showing estimated tumour purity of non-recurrent (blue) and recurrent (red) ccRCC tumours by xCell using gene expression data from PCS and DRS. **E.** Grouped dot plot showing relative population of $CD8^+$ T cells within immune infiltrates of non-recurrent (blue) and recurrent (red) ccRCC tumours estimated by CIBERSORTx using DRS gene expression data. **F.** Correlation between CIBERSORTx estimated $CD8^+$ T cells fraction amongst immune infiltrates in non-recurrent (blue) and recurrent (red) ccRCC tumours, generated by PCS and DRS gene expression data. **G.** Grouped dot plot showing relative cell fraction of $CD8^+$ T cells in non-recurrent (blue) and recurrent (red) ccRCC tumours estimated by EPIC using PCS and DRS gene expression data. **H.** Grouped dot plot showing *NOS3* and *CCL5* gene expression (Reads per million (RPM)) in non-recurrent (blue) and recurrent (red) ccRCC PCS gene expression data. For **C** and **F**, $R^2$ values were computed to measure goodness-of-fit and p values were generated from F-test, with $p \leq 0.05$ considered statistically significant. For **A**, **B**, **D**, **E**, **G** and **H**, two-tailed Mann-Whitney U tests were used with $p \leq 0.05$ considered significant. * = $p < 0.05$, ** = $p < 0.01$, **** = $p < 0.0001$. p value of non-significant results is indicated in graph. Centre line represents median for each group.

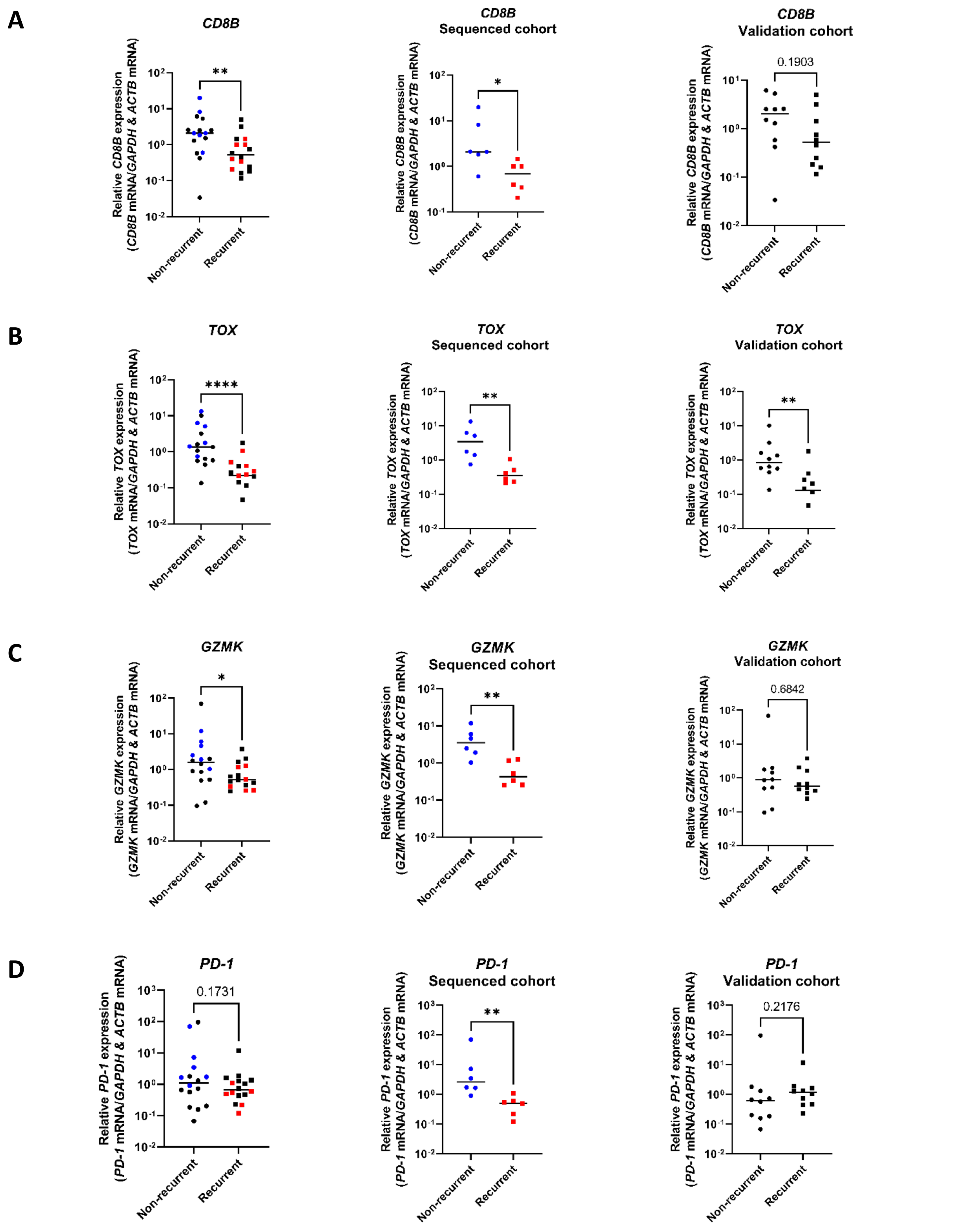


**Figure S5: Validation of sequencing results via qRT-PCR**. **A.** *CD8B*, **B.** *TOX*, **C.** *GZMK* and **D.** *PD-1* mRNA levels measured by qRT-PCR in recurrent and non-recurrent tumours from sequenced cohort (blue and red, middle, n = 12) and validation cohort (black, right, n = 20) relative to average mRNA levels in non-recurrent tumours. mRNA levels were normalised to *GAPDH* and *ACTB*. Plots showing data from both sequenced cohort and validation cohort (left) replicate content of Fig. 2 from the main body of the paper to provide a clearer visual representation of data. two-tailed Mann-Whitney U tests were used with $p \leq 0.05$ considered significant. * = $p < 0.05$, ** = $p < 0.01$, **** = $p < 0.0001$. p value of non-significant results is indicated in graph. Centre line represents median for each group.

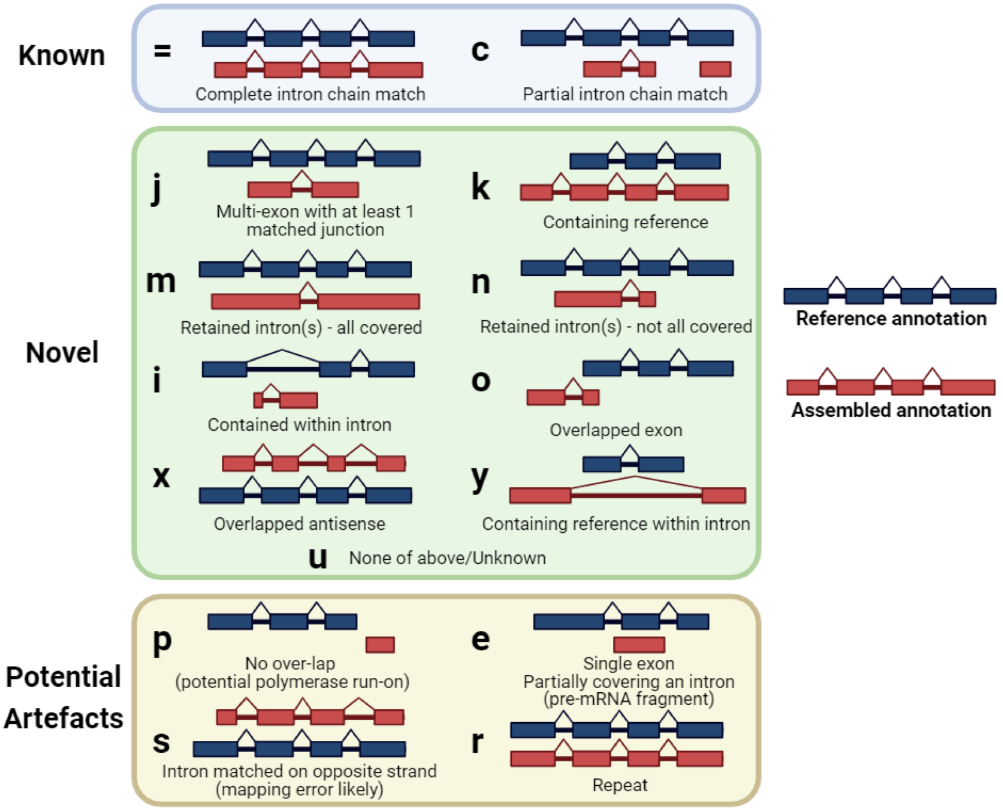


**Figure S6: Graphical representation of GffCompare transcript class codes.** Transcript class codes were categorised into 'Known' ('=' : Complete intron chain match, 'c': Partial intron chain match), 'Novel' ('j' : Multi-exon with at least 1 matched junction', 'k' : Containing reference, 'm' : Retained intron(s) – all covered, 'n' : Retained intron(s) – not all covered, 'i' : Contained within intron, 'o' : Overlapped exon, 'x' : Overlapped antisense, 'y' : Containing reference within intron, 'u' : None of above/Unknown), and 'Potential Artefacts' ('p' : No over-lap, 'e' : Single exon partially covering an intron, 's' :Intron matched on opposite strand, 'r' : Repeat).

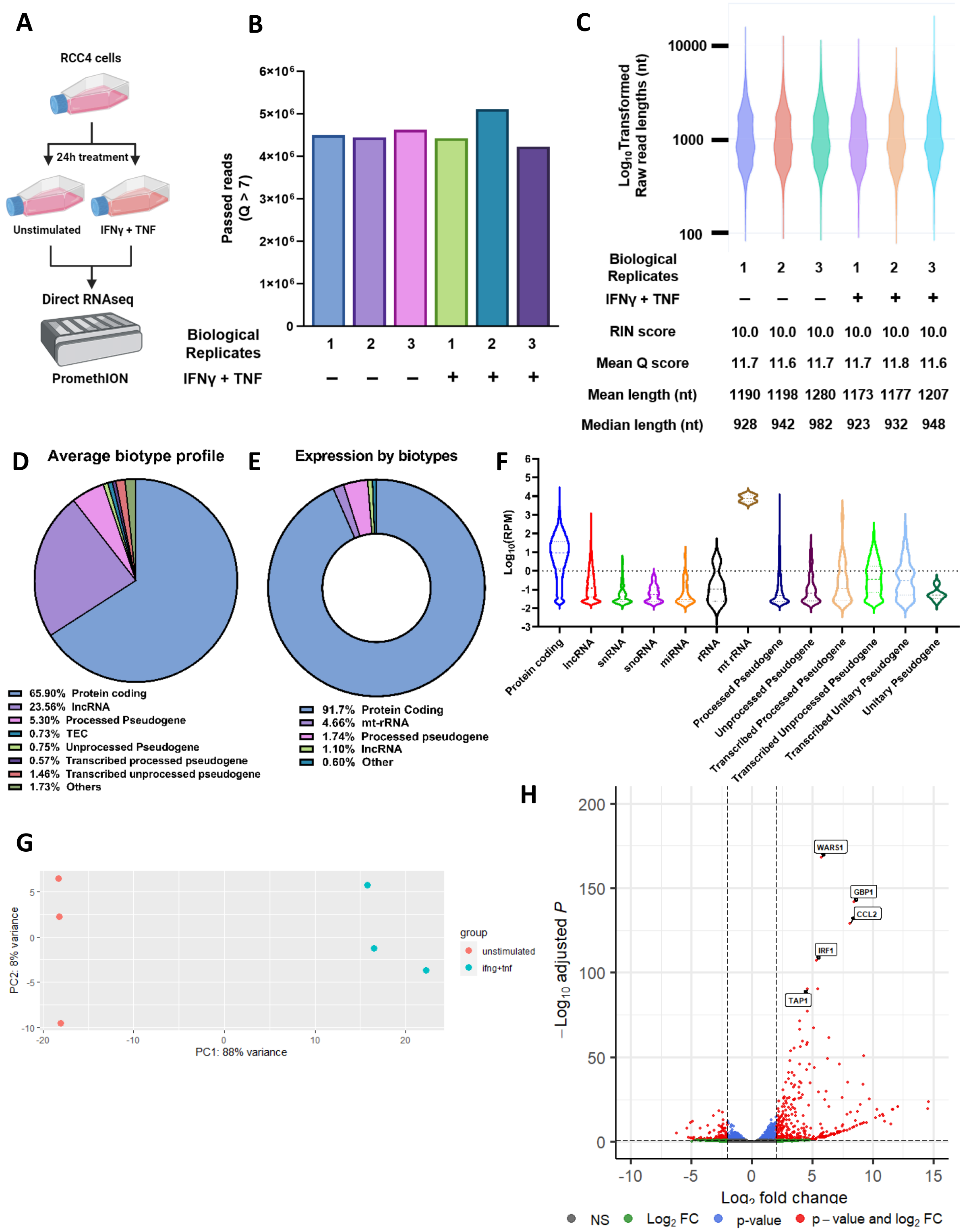


**Figure S7: Sequencing statistics of IFNγ + TNF treated and untreated RCC4 cells. A** Summary workflow for DRS of RCC4 cells. Figure made with Biorender. **B.** Bar chart showing the number of sequencing reads generated by DRS of RCC4 cells that passed the quality filter (Q score >7). **C.** Violin plot showing $Log_{10}$ transformed raw read lengths of passed reads generated by DRS of RCC4 cells. RIN score, mean read Q score, mean and median read length for each sequencing dataset are listed in the table below violin graph. **D.** Pie chart depicting the average proportions of RNA biotypes of genes mapped by DRS of RCC4 cells.

**E.** Pie chart showing the average proportions of RNA biotypes of mapped genes by expression levels from DRS of RCC4 cells. **F.** Violin plot depicting the distribution of gene expression levels ($Log_{10}$RPM) of mapped genes by biotypes from DRS of RCC4 cells, with first, third quartiles and median shown as horizontal line within each plot. **G.** DESeq2 generated PCA plots using expression data from DRS of RCC4 cells, showing PCA of unstimulated and IFNγ+TNF treated groups. **H.** Volcano plot showing differentially expression genes (red) between unstimulated and IFNγ+TNF treated RCC4 cells from DRS data. Dotted lines indicate significance threshold ($|log_2FoldChange| \geq 2$, $p_{adj} \leq 0.1$). Names of top 5 most significantly differentially expressed genes (by $p_{adj}$) are shown.

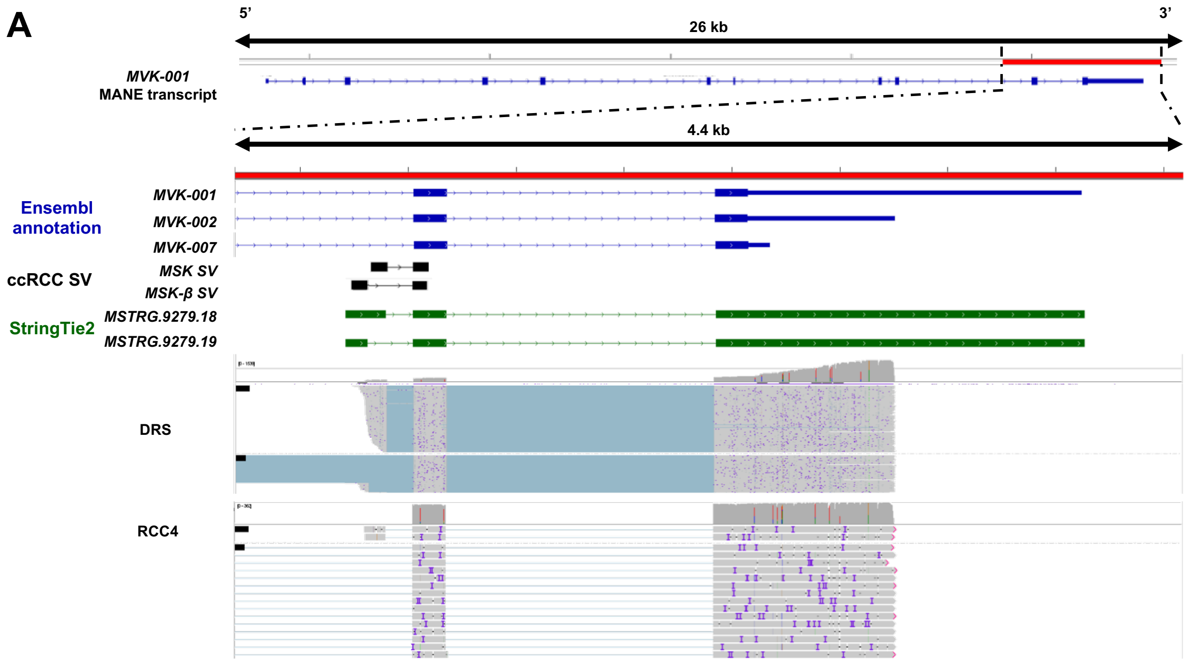


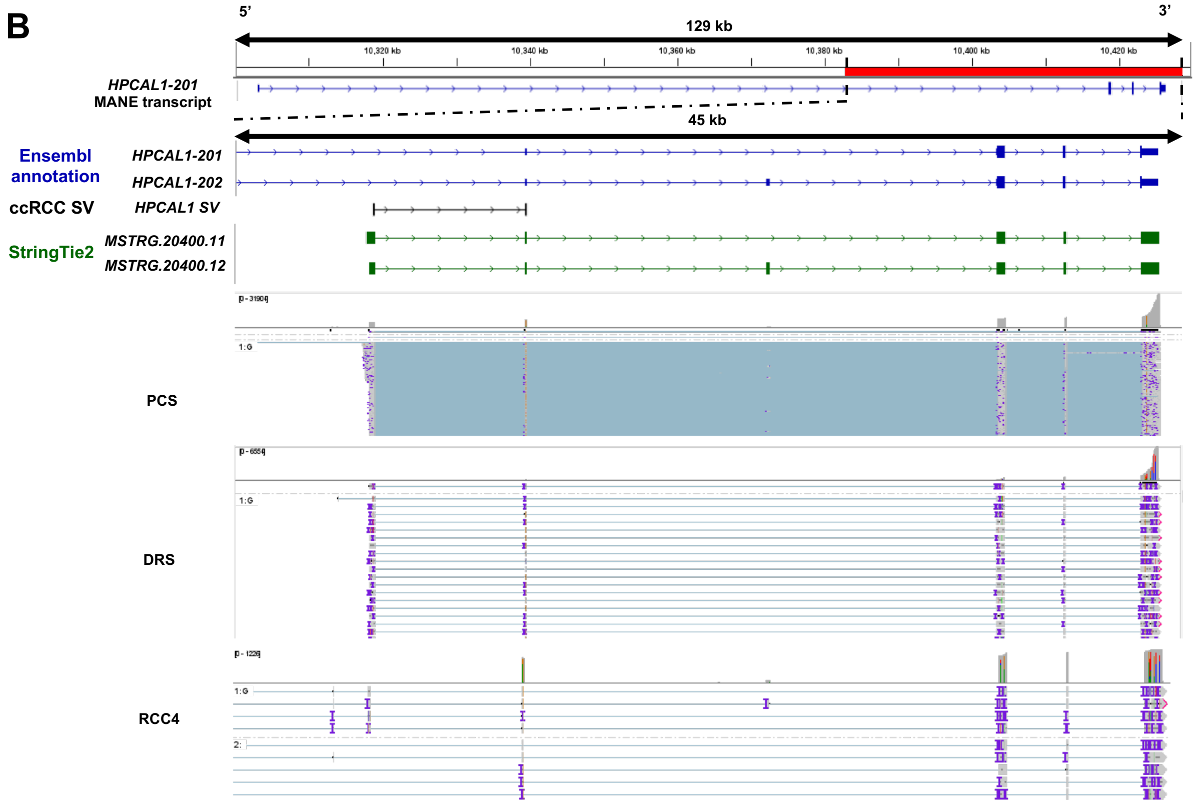


**Figure S8: Visualisation of long-read RNAseq reads mapping to ccRCC-specific splice junctions. A.** IGV visualisation of *MVK* reference annotations (blue), ccRCC specific *MVK* splice junctions (black), Stringtie2 assembled novel transcripts (green), DRS of ccRCC tumour samples (labelled as DRS) and RCC4 coverage tracks (grey) and sequencing reads aligned to the reference genome in the region of interest (red bar, hg38 chr12:109,594,200 – 109,598,600). **B.** IGV visualisation of *HPCAL1* reference annotations (blue), ccRCC specific *MVK* splice junctions (black), Stringtie2 assembled novel transcripts (green), PCS and DRS of ccRCC tumour samples (labelled as DRS) and RCC4 coverage tracks (grey) and sequencing reads aligned to the reference genome in the region of interest (red bar, hg38 chr2:10,300,100-10,429,600).

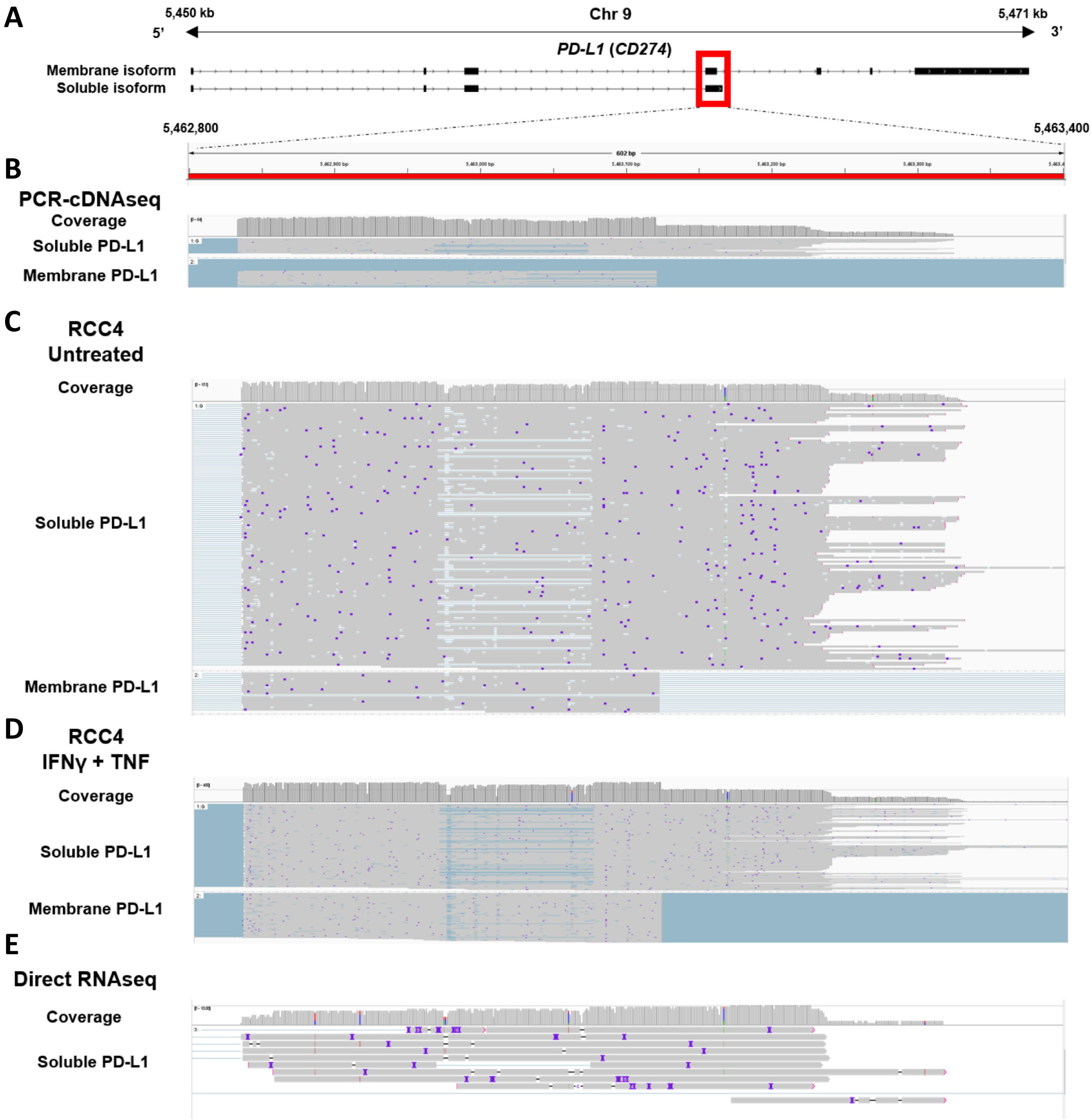


**Figure S9: Sequencing reads aligned to *PD-L1* exon 4. A.** IGV visualisation of reference annotation of *mPD-L1* isoform (ENST00000381577) and *sPD-L1* (black, NM_001314029). **B.** ccRCC tumours PCR-cDNA sequencing coverage plot and reads aligned to *PD-L1* exon 4 region (chr9:5,462,800 – 5,463,400). **C.** Untreated RCC4 DRS coverage plot and reads aligned to *PD-L1* exon 4 region (chr9:5,462,800 – 5,463,400). **D.** IFNγ + TNF treated RCC4 DRS coverage plot and reads aligned to *PD-L1* exon 4 region (chr9:5,462,800 – 5,463,400). **E.** ccRCC tumours DRS sequencing coverage plot and reads aligned to *PD-L1* exon 4 region (chr9:5,462,800 – 5,463,400).

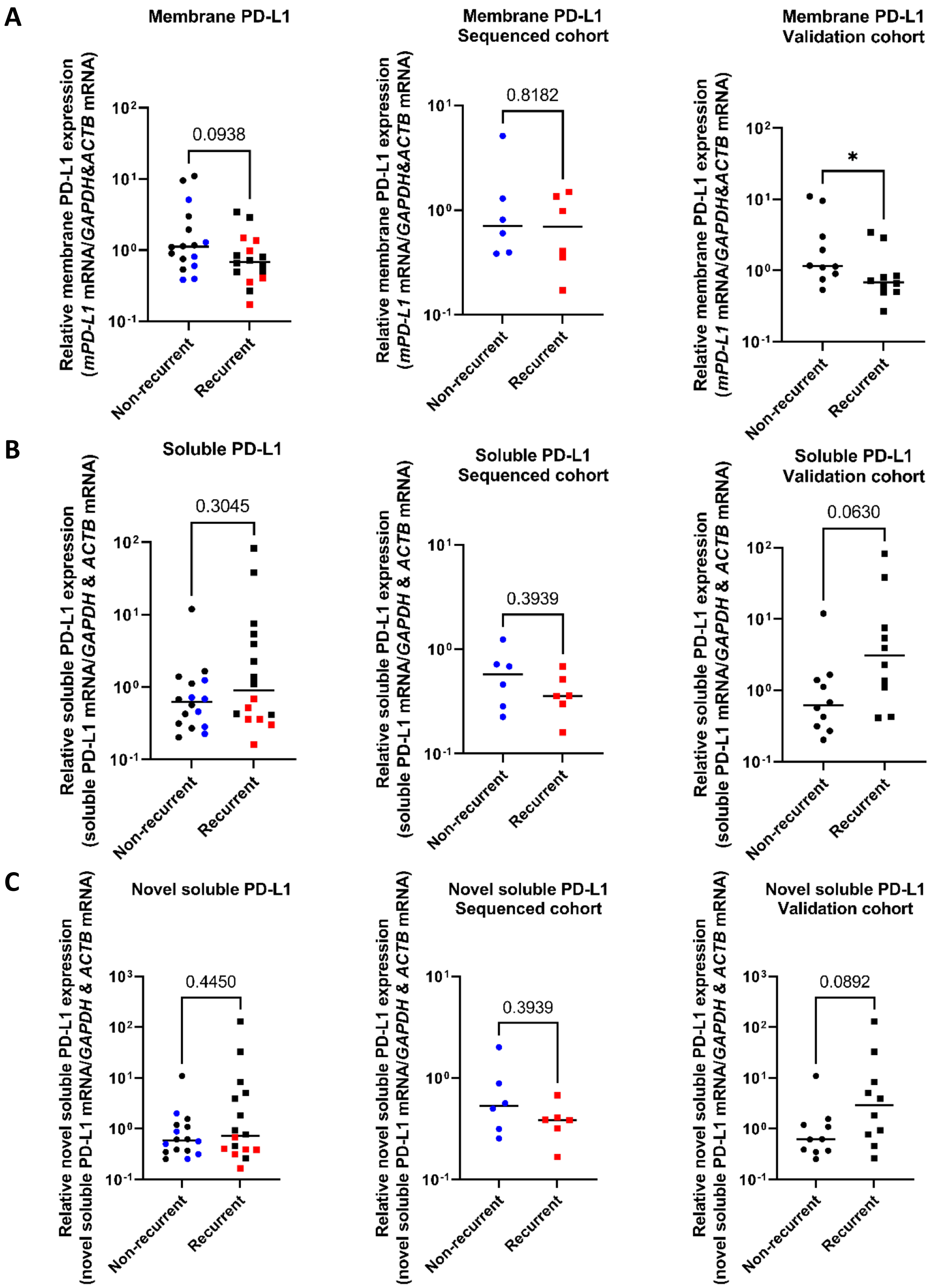


**Figure S10: Validation of *PD-L1* sequencing results and novel *sPD-L1* isoform via qRT-PCR**. **A.** *mPD-L1*, **B.** *sPD-L1* and **C.** Novel *sPD-L1* mRNA levels measured by qRT-PCR in recurrent and non-recurrent tumours from sequenced cohort (blue and red, middle, n = 12) and validation cohort (black, right, n = 20) relative to average mRNA levels in non-recurrent tumours. mRNA levels were normalised to *GAPDH* and *ACTB*. Plots showing data from both sequenced cohort and validation cohort (left) replicate content of Fig. 5 from the main body of the paper to provide a clearer visual representation of data. Two-tailed Mann-Whitney U tests were used with p ≤ 0.05 considered significant. * = $p < 0.05$, p value of non-significant results is indicated in graph. Centre line represents median for each group.

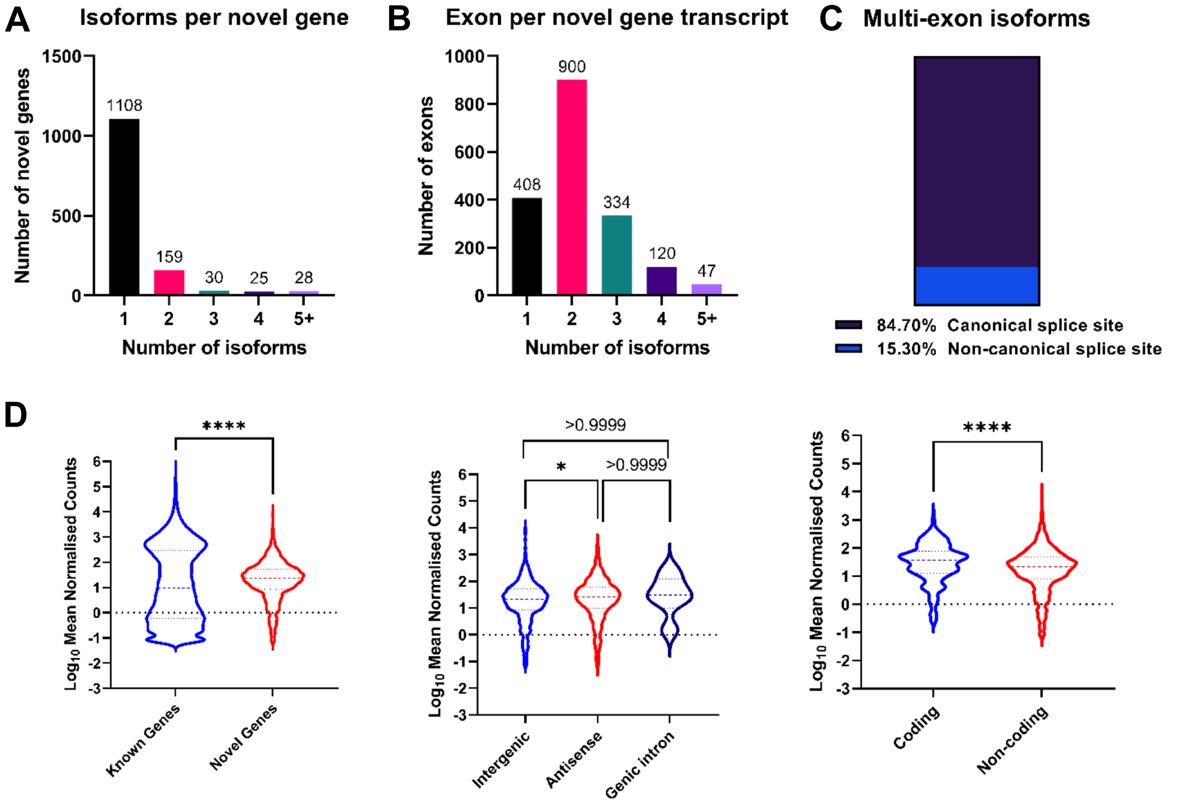


E

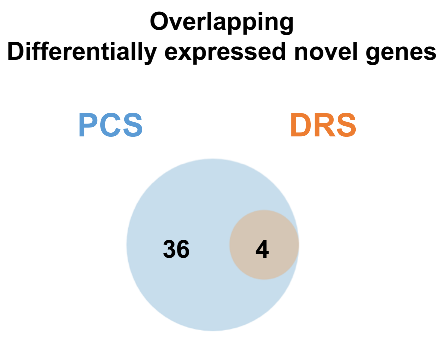


| | PCS | | DRS | |
|---|---|---|---|---|
| Novel Genes | $Log_2FC$ | $p_{adj}$ | $Log_2FC$ | $p_{adj}$ |
| MSTRG.1938 | -2.550 | 0.055 | -2.989 | 0.031 |
| MSTRG.21132 | 5.227 | $1.88 \times 10^{-7}$ | 5.994 | 0.035 |
| MSTRG.29728 | 2.951 | 0.051 | 3.441 | 0.067 |
| MSTRG.38727 | 4.821 | 0.099 | 5.099 | 0.092 |

F

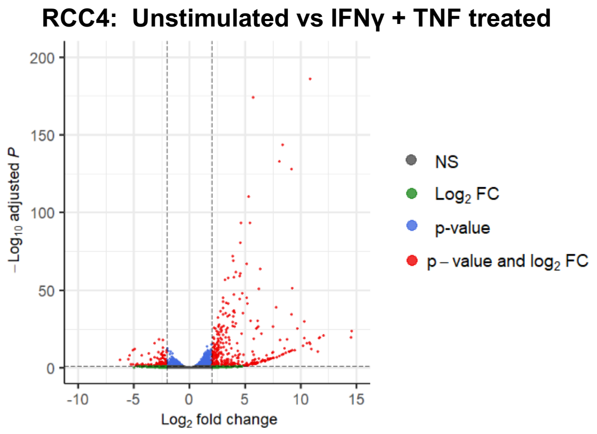


| | Down regulated | Up regulated |
|---|---|---|
| Known | 111 | 324 |
| Novel | 4 | 9 |

**Figure S11: Characterisation of novel genes. A.** Bar chart showing the distribution of the number of isoforms per PCS mapped novel gene (n = 1,350). **B.** Bar chart showing the distribution of the number of exons per isoform from mapped novel genes (n = 1,350). **C.** Bar chart showing the proportion of multi-exon novel gene isoform (n = 758) that consists of canonical splice sites (purple) or non-canonical splice sites (blue). **D.** Violin plots depicting the distribution of gene expression levels ($Log_{10}RPM$) of novel genes versus known genes (Ensembl annotation 105); Novel genes that are classified by SQANTI3 as intergenic versus antisense and genic intron; Novel genes that are classified as coding versus non-coding.Two-tailed Mann-Whitney U tests (left, right) and Kruskal-Wallis test (centre) were used with $p \leq 0.05$ considered significant. **** = $p < 0.0001$, * = $p < 0.05$. p value of non-significant results is indicated in graph. **E.** Venn diagram showing the number of overlapping differentially expressed novel genes between PCS and DRS of ccRCC tumour samples. $Log_2$FoldChanges and $p_{adj}$ values of the overlapping differentially expressed novel genes by DESeq2 analysis are shown in table below the diagram. **F.** Volcano plots showing differentially expressed genes (red) between unstimulated and IFNγ+TNF treated RCC4 cells using Stringtie assembled reference. Number of differentially expressed novel and known genes are shown in table below plots.

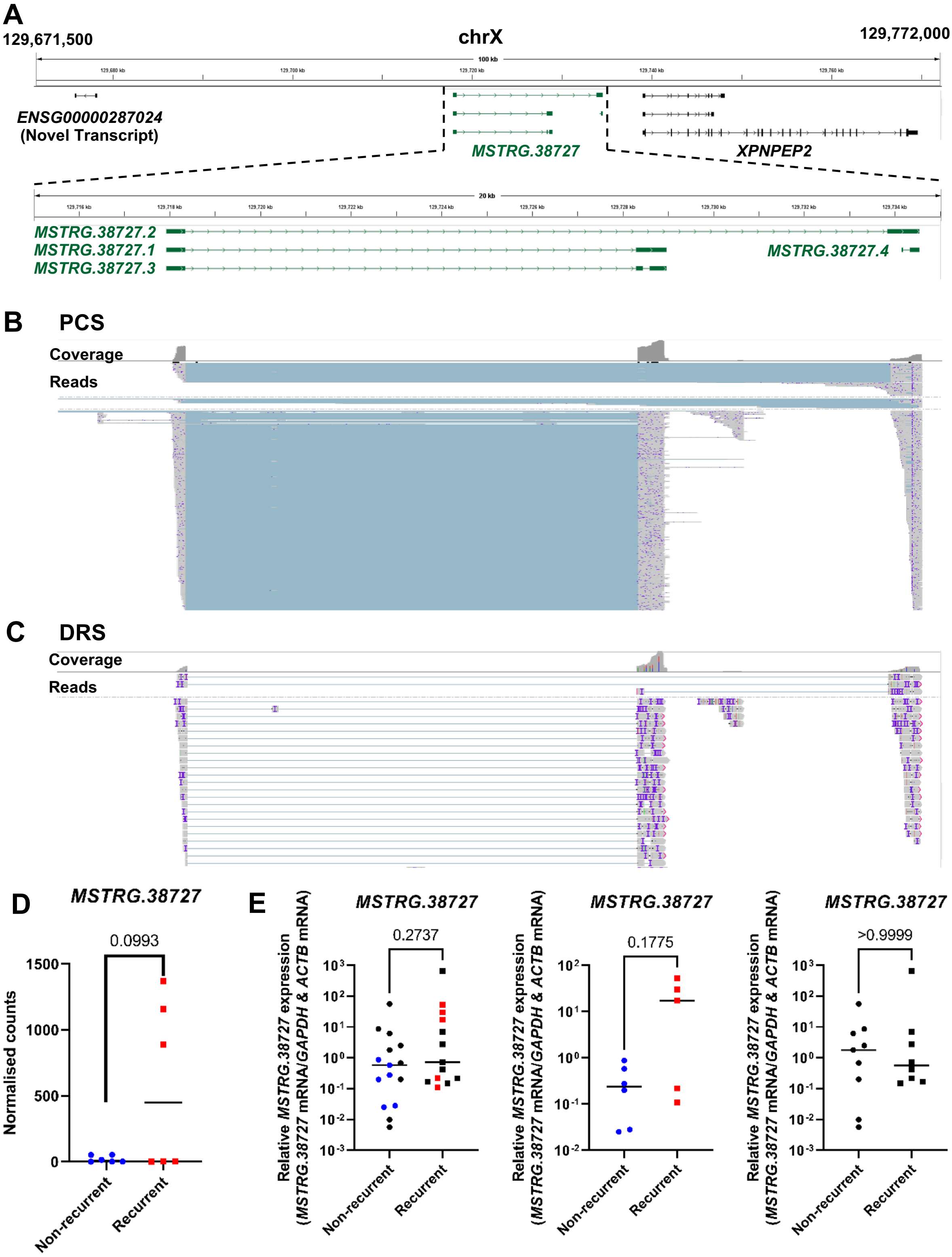


**Figure S12: Visualisation and expression validation of novel gene *MSTRG.38727*. A.** IGV visualisation of *MSTRG.38727* (green) and the closest neighbouring genes (*ENSG00000287024 (novel transcript)* and *XPNPEP2*) in the Ensembl reference annotation (Ensembl release 105) at chrX:129,671,500 – 129,772,000 (Top tracks); *MSTRG.38727* isoforms structures (*MSTRG.38727.1*, *MSTRG.38727.2*, *MSTRG.38727.3*, *MSTRG.38727.4*) at chrX: 129,715,000 – 129,735,000 (Green, bottom tracks). **B.** IGV coverage track and reference genome aligned reads from PCS of ccRCC tumour samples in the region of interest (chrX: 129,715,000 – 129,735,000). **C.** IGV coverage track and reference genome aligned reads from PCS of ccRCC tumour samples in the region of interest (chrX: 129,715,000 – 129,735,000). **D.** Grouped dot plot showing reference DESeq2 normalised *MSTRG.38727* expression in non-recurrent (blue) and recurrent (red) tumours' PCS data. DESeq2 $p_{adj}$ value is shown in graph. Centre line represents median for each group. **E.** *MSTRG.38727* mRNA levels measured by qRT-PCR in recurrent and non-recurrent tumours from sequenced cohort (blue and red, middle, n = 12), validation cohort (black, right, n = 20) and both cohorts relative to average mRNA levels in non-recurrent tumours. mRNA levels were normalised to *GAPDH* and *ACTB*. Two-tailed Mann-Whitney U tests were used with $p \leq 0.05$ considered significant. p value of non-significant results is indicated in graph. Centre line represents median for each group.
